# Phenotypic illusion but genetic decoupling between osteoarthritis and Parkinson’s disease

**DOI:** 10.64898/2026.09.05.26362301

**Authors:** Shenxu Yu, Zhaoyang Wu, Huaikang Zhou, Peijian Tong

## Abstract

Observational studies have repeatedly linked osteoarthritis (OA) with Parkinson’s disease (PD), but whether this reflects causality, shared genetic architecture, or artefactual mechanisms remains unresolved. We triangulated six layers of evidence: two population cohorts, genome-wide and local genetic architecture, bidirectional Mendelian randomization, shared-locus mapping, cell-type-specific analyses, and single-cell expression validation. In CHARLS (n = 7,587), knee OA was associated with higher PD risk (OR 1.88, 95% CI 1.24–2.84) with consistent depression-mediated attenuation (≈34%). In NHANES 2011–2018, the association was positive but imprecise (OR 1.94, 95% CI 0.44–8.52) and non-specific. Genetically, OA and PD were decoupled: genetic correlation was null-to-negative, no local correlation survived correction, and bidirectional MR was null across all pre-specified chains and sensitivity analyses. Conditional FDR identified 732 independent shared loci without directional bias; colocalization converged on a single robust shared region at MAPT 17q21.31 (posterior probability 0.975). Cell-type-specific and single-cell expression analyses did not support an expression-mediated pathway. The OA– PD association is therefore largely consistent with a phenotypic illusion (reproducible but proxy-sensitive and depression-mediated), with genome-wide genetic decoupling and only locus-restricted sharing at 17q21.31.

## Introduction

Osteoarthritis (OA) and Parkinson’s disease (PD) are chronic, age-related conditions with rapidly rising global burdens. OA is the most common form of arthritis: 595 million people (95% uncertainty interval [UI] 535–656), representing 7.6% of the global population, were affected in 2020 (a 132% increase since 1990), with knee OA the most common site and knee cases projected to rise by a further 74.9% (95% UI 59.4–89.9) by 2050^1^. OA was also the seventh-ranked cause of years lived with disability (YLDs) among adults aged ≥70 years^1^. PD is an age-dependent neurodegenerative disorder: global prevalence more than doubled between 1990 and 2016, from 2.5 to 6.1 million cases, and PD was responsible for 211,296 deaths (95% UI 167,771–265,160) in 2016^2^. By 2050, 25.2 million (95% UI 21.7–30.1) people are projected to be living with PD, a 112% increase from 2021 driven predominantly by population ageing, with East Asia alone accounting for an estimated 10.9 million cases^3^. Nervous-system disorders as a group were the leading cause of disability-adjusted life-years (DALYs) globally in 2021, contributing 443 million DALYs^4^. The parallel rise of these two conditions raises the question of whether joint degeneration and neurodegeneration are connected beyond demographic co-occurrence.

Against this shared ageing backdrop, OA, particularly knee OA, has been repeatedly linked to higher PD risk in population-based studies across health systems and designs: a Danish cohort (slightly elevated standardized incidence ratio)^5^, a Taiwanese longitudinal study^6^, US National Health and Nutrition Examination Survey (NHANES) cross-sections^7^, and a UK Biobank study of age-related musculoskeletal diseases^8^. Effects are modest (relative risks ≈1.1–2.0) and directionally consistent, but the mechanistic basis remains unresolved.

The association could reflect genuine causality (chronic low-grade inflammation, shared ageing biology, or the musculoskeletal–brain axis) or arise from symptom–behaviour pathways (pain → depression → reduced physical activity → increased health-care contact), proxy-outcome misclassification (e.g., restless-legs-syndrome medications counted as anti-Parkinson drugs), or residual confounding. Whether knee OA is a modifiable PD risk factor therefore remains an open question with direct implications for prevention.

Genetics offers a confounding-robust test of causality, yet the MR literature is contradictory. Early bidirectional analyses reported a positive OA-to-PD effect^9^; a 2026 NHANES-plus-GWAS analysis concluded that genetic predisposition to knee OA increased PD risk (OR 1.49, 95% CI 1.11–2.02)^10^; and a multi-level arthritis–neurodegeneration study reported OA-associated risks of Alzheimer disease (AD), PD, and autonomic-nervous-system disease, RA-associated AD risk without an RA–PD association, and context-dependent in vivo RNF40 function^11^.

No published study has systematically characterised the OA–PD relationship at the level of genome-wide genetic correlation, local genetic architecture, conditional-FDR shared-locus structure, or OA-exposure-driven cell-type-specific causality. Positive MR claims in this area typically lack sample-overlap auditing, proxy-case sensitivity, colocalization, and cell-level validation (now standard in rigorous genetic epidemiology), so conflicting results may reflect instrument selection, sample overlap, or proxy outcome definitions rather than true causality.

Even if genetically decoupled genome-wide, limited shared regulatory signals could act through specific brain cell types. Cell-type-specific cis-eQTL maps (eight brain cell types, 192 donors; Bryois et al. 2022)^12^ enable exposure-side cell-type-specific MR (csMR) for OA loci. A 2026 Movement Disorders study demonstrated cell-type-specific causal associations for ARL17A, ARL17B, KANSL1, and LRRC37A across seven brain cell types^13^; it established the methodological framework and drew attention to MAPT 17q21, but considered PD risk loci only. Three PD midbrain single-nucleus RNA-seq datasets (Smajić 2022^14^; Kamath 2022^15^; Martirosyan 2024^16^) provide the expression-level data needed to test direction consistency.

Here we report a six-layer triangulation of the OA–PD association: (i) phenotypic analyses in two cohorts with temporal depression mediation, competing-risk, and proxy-definition sensitivity; (ii) genome-wide and local genetic correlation; (iii) bidirectional two-sample MR under strict no-overlap and no-proxy-PD conditions, including a direct instrument-set audit of the positive claim^10^; (iv) conditional/conjunctional FDR, SuSiE colocalization^17^, and SMR/HEIDI for shared-locus mapping; (v) cell-type-specific MR and single-cell enrichment; and (vi) expression validation in three PD midbrain snRNA-seq datasets. We hypothesized a “phenotypic illusion” (reproducible but non-specific, proxy-sensitive, and substantially mediated by symptom/behaviour pathways), with genome-wide “decoupling” and at most limited, direction-unbiased sharing at MAPT 17q21.31. An OA × AD contrast tested whether the decoupling signature is disease-specific. All definitions, thresholds, and decision rules were pre-specified in the analysis protocol, which was archived in the OSF project (https://osf.io/rgeaf). Because analyses had begun before registration, the OSF registration (completed 25 August 2026) is retrospective; amendments are documented (Supplementary Note N4).

## Results

### Phenotypic association

In CHARLS (2011–2020; ≥50 years, no baseline stroke or memory-related disease), baseline knee OA predicted higher 2020 self-reported PD (the primary adjusted model; n = 7,587, events = 103; OR 1.88, 95% CI 1.24–2.84, p = 0.0028) and persisted under inverse-probability weighting for attrition (OR 2.49, 95% CI 1.61–3.85) and with death as a competing event (Fine-Gray SHR 2.69, 95% CI 1.75–4.13; 9-year cumulative incidence 0.89% vs 2.37% without vs with knee OA). Sensitivity analyses across pre-specified exposure definitions and samples are shown in Supplementary Fig. 2. In the pre-specified temporal mediation design (2011 exposure → 2013/2015 depression → 2020 PD; n = 8,486), the total indirect log-OR was 0.192 (95% CI 0.042–0.355; ≈23.5% of the total effect), and adding depression attenuated the OR by ≈34% (1.88 → 1.52, p = 0.052)^18^. A multi-mediator decomposition (depression and incident stroke; n = 7,587) confirmed independent indirect effects (total indirect logOR 0.226, 95% CI 0.089–0.365; depression-specific 0.184, 0.045–0.307; stroke-specific 0.342, 0.090–0.653; proportion mediated 35.8%), and the temporal sequential decomposition (n = 6,360) remained directionally consistent (total indirect logOR 0.146; IPW-attenuated to 0.134). The decomposition designs therefore converge on depression as a phenotypic pathway.

In NHANES 2011–2018, physician-diagnosed OA (any site; no site-specific information) was assessed against a PD-specific prescription outcome (levodopa, MAO-B, or COMT inhibitors, excluding restless-legs-syndrome agonists; n = 6,320, events = 20; complete-case mediation sample n = 6,234, events = 19). The total association was positive but imprecise in this cross-sectional sample (survey-weighted OR 1.94, 95% CI 0.44–8.52, p = 0.38; Firth-penalized OR 2.47, 95% CI 0.96–6.43, p = 0.062). Sensitivity analyses across the ten pre-specified scenarios are shown in Supplementary Fig. 3. The PHQ-9-defined depression indirect effect was significant and directionally consistent with CHARLS (0.201, 95% CI 0.116–0.321). As a cross-sectional sensitivity analysis (mediators measured concurrently, unlike the CHARLS temporal design), adding depression as a covariate attenuated the total OR by ≈60% on the log-odds scale (1.94 → 1.30); this add-mediator coefficient change is distinct from the formal mediation indirect effect of 0.498 reported in Table 2. Given the few PD-specific events, specificity was evaluated across proxy and negative-control outcomes: OA was associated with restless-legs-syndrome medication (OR 3.48), asthma (OR 1.91), and hearing impairment (OR 1.59), whereas the PD-specific outcome was not significant, indicating that the “PD” association is non-specific to OA and sensitive to outcome definition; negative-control associations reflect health-care-contact and comorbidity-related selection rather than PD-specific biology. In the extended mediator screen (sleep, obesity, physical inactivity, cardiovascular comorbidity), depression was the only significant mediator (FDR<0.001); the negative cardiovascular-comorbidity estimate was treated as a small-sample artefact.

The OA–PD association therefore reproduces longitudinally in CHARLS and directionally in NHANES. It is substantially mediated by depression and is non-specific and proxy-sensitive. These features are consistent with a phenotypic illusion rather than an OA-specific causal effect (Figs. 1–2; Tables 1–2; Supplementary Tables S2–S3). Forest-plot annotation conventions used throughout the manuscript are described in Supplementary Note N6.

**Fig. 1.**
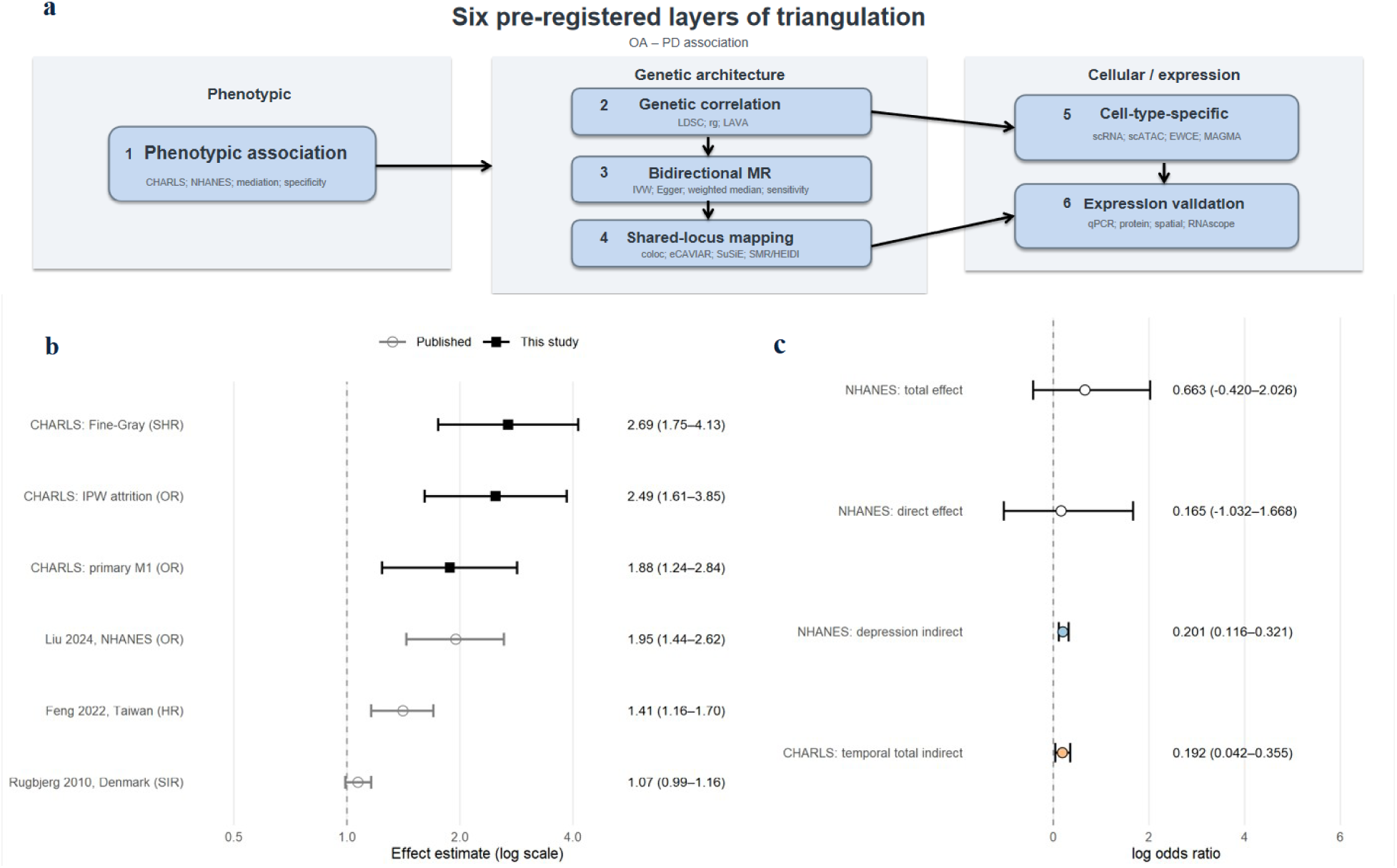
Study design and phenotypic layer. (a) Six-layer triangulation framework (phenotypic → genetic correlation → MR → shared-locus mapping → cell-type-specific → expression validation; OA × AD contrast shown). (b) CHARLS effect-size forest plot (primary model, IPW, Fine-Gray) alongside published estimates (Feng 2022; Liu 2024; Rugbjerg 2010). (c) Depression mediation decomposition (NHANES indirect effect 0.201, 95% CI 0.116–0.321; CHARLS temporal total indirect logOR 0.192, 95% CI 0.042–0.355; CHARLS sequential total indirect logOR 0.146 (IPW-attenuated to 0.134)). Filled points: light blue = NHANES depression-mediated indirect effect, light orange = CHARLS temporal total indirect effect, open = total/direct effects.

**Fig. 2.**
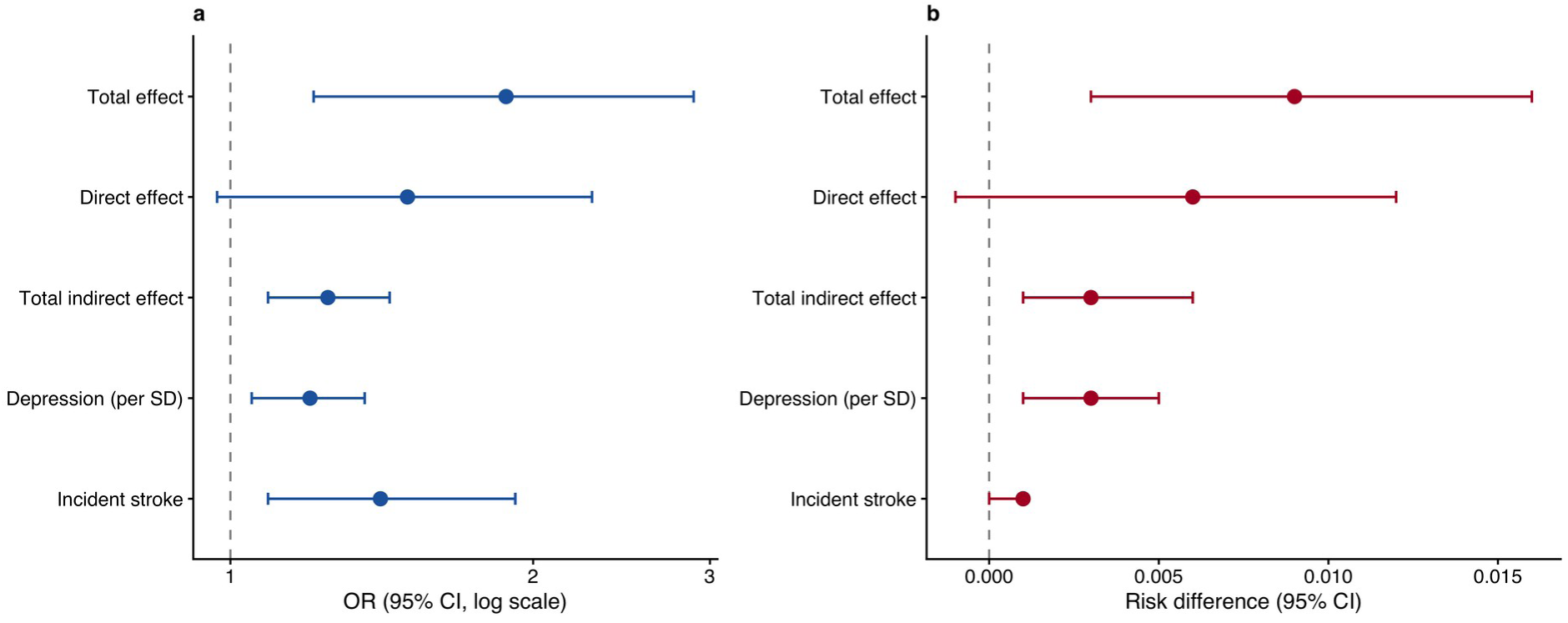
CHARLS multi-mediator decomposition (n = 7,587). Total, direct, and indirect effects of knee OA on PD through depression and incident stroke on the OR scale (a) and risk-difference scale (b); proportion mediated 35.8%.

**Table 1.**
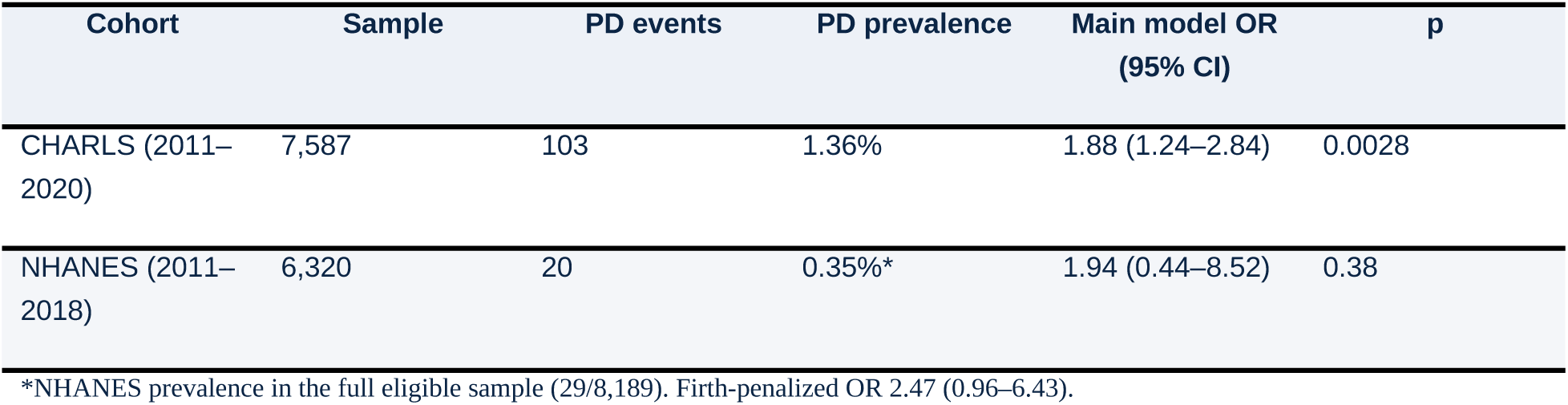
Cohort characteristics and main phenotypic association.

**Table 2.**
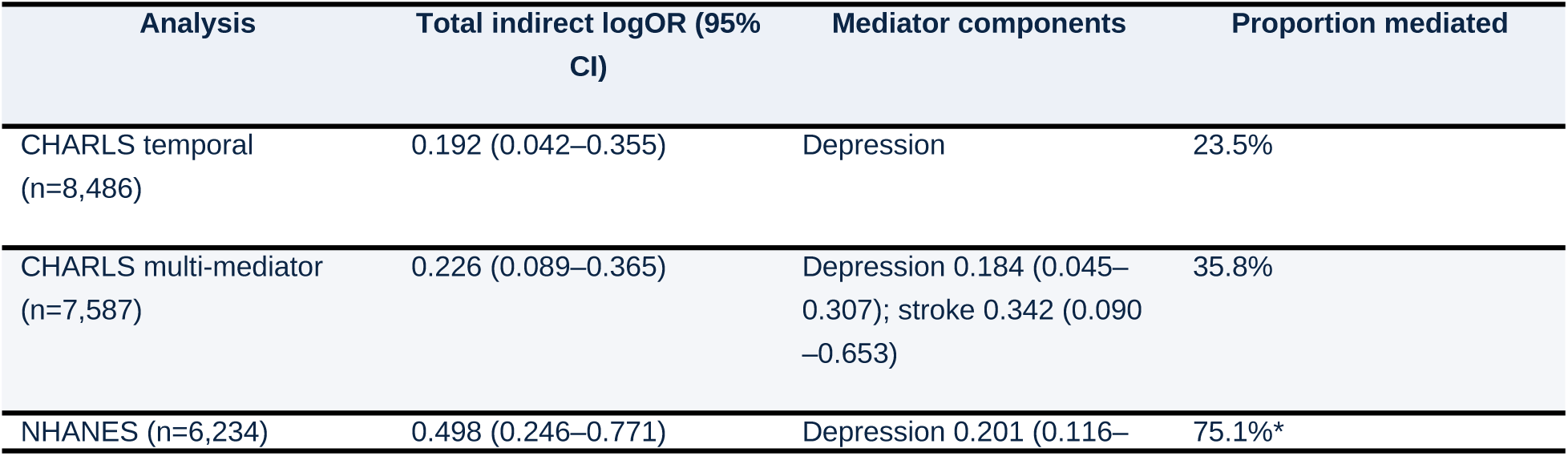
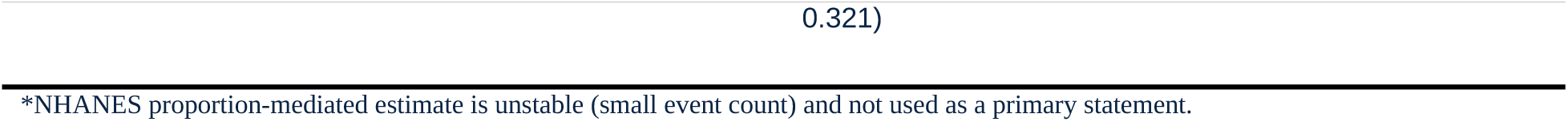
Depression and comorbidity mediation decomposition.

### Genome-wide genetic correlation

LD score regression^19^ estimated genome-wide genetic correlation between OA and PD across all pre-specified combinations (Fig. 3; Table 3; Supplementary Table S4). In combination A (FinnGen OA × Nalls 2019 PD^20^; no sample overlap), genetic correlation was negative for all-site OA (rg = −0.091, SE 0.034, p = 0.008; p = 0.213 after Bonferroni) and for hip (rg = −0.085) and knee (rg = −0.072) subtypes (nominal p = 0.008–0.038). Combination B (Hatzikotoulas 2025 EUR OA^21^ × Nalls 2019 PD^20^; maximum power, UK Biobank overlap quantified) gave null-to-negative estimates (all-site rg = −0.043, p = 0.16), as did the internal combination C (FinnGen OA × FinnGen PD), the no-proxy combination E (FinnGen/Hatzikotoulas OA × IPDGC 2021 no-UKBB^22^), the trans-ancestry combination F (Kim 2024 GP2^23^), and total knee replacement as a hard knee-OA outcome (rg = −0.053). No combination yielded a significant positive genetic correlation.

**Fig. 3.**
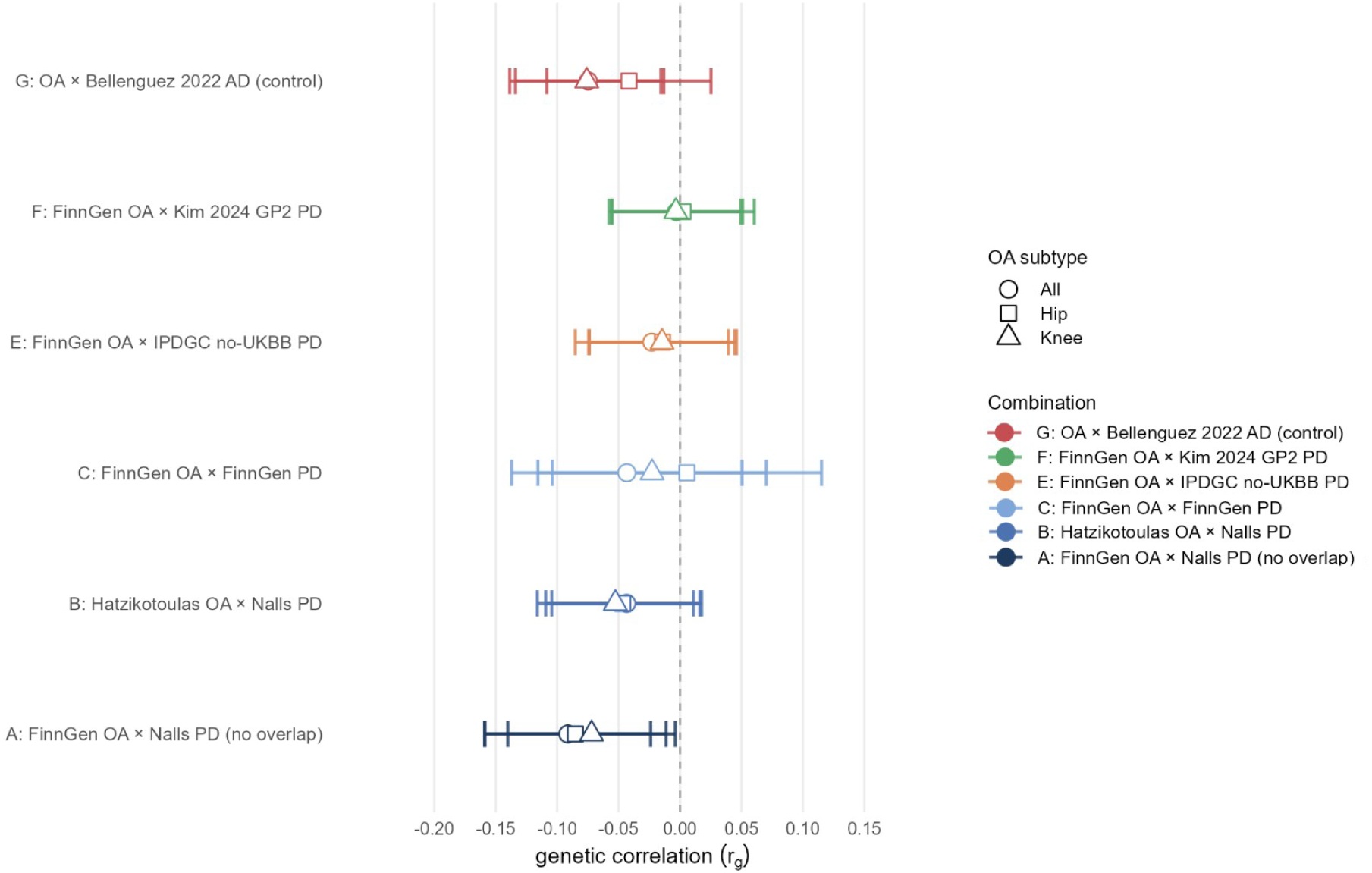
Genome-wide genetic correlation. LDSC genetic-correlation forest plot across pre-specified combinations (OA × PD: A/B/C/E/F; OA × AD: G; knee/hip/all subtypes). Filled points = Bonferroni-significant; open circles = gcov-intercept-constrained sensitivity; dashed line = null (rg = 0). LAVA local-rho distributions are reported in Supplementary Table S4.

**Table 3.**
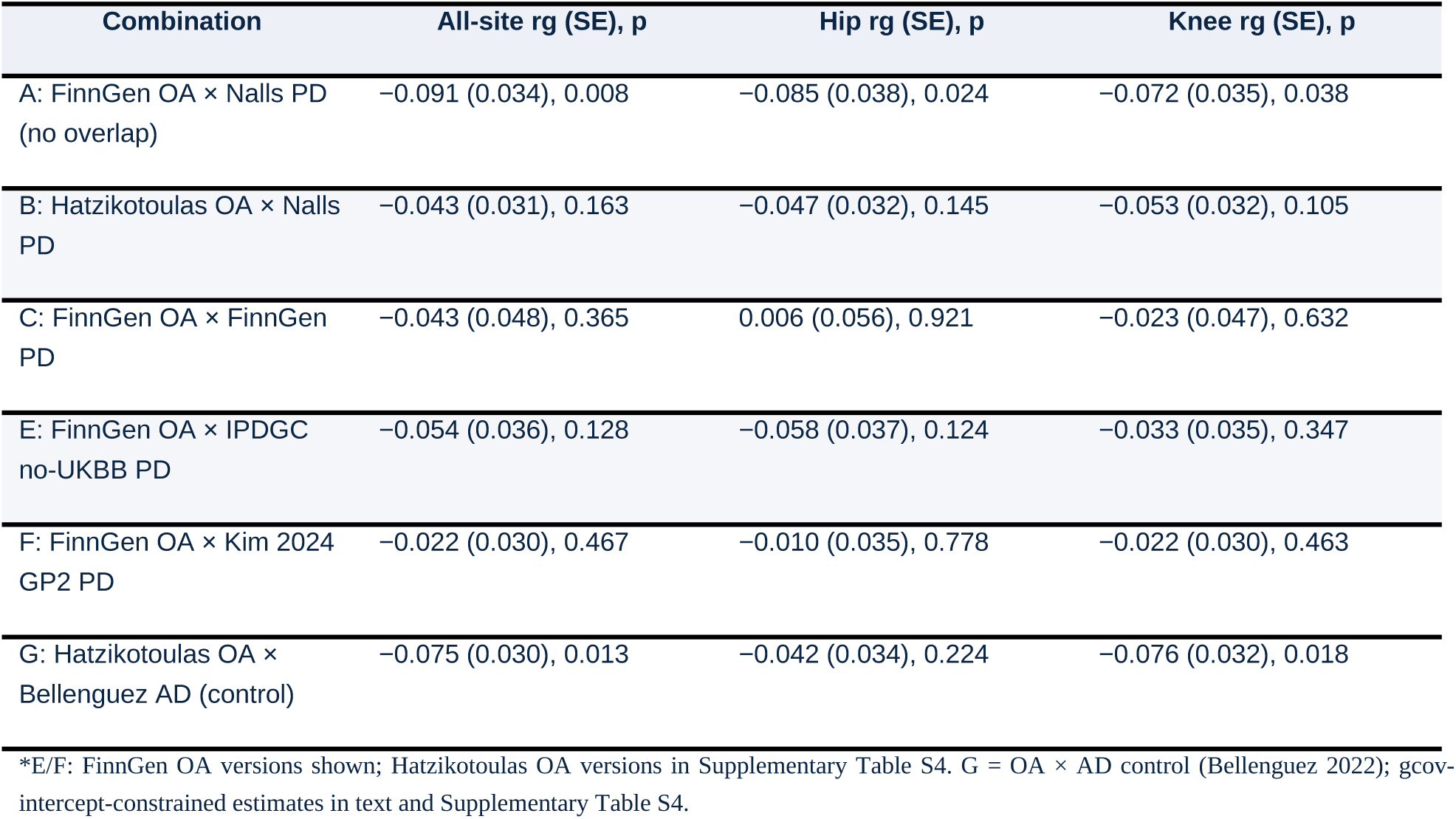
Genome-wide genetic correlation (LDSC rg) across pre-specified combinations.

LAVA across 2,495 genomic blocks found no region surviving correction in any combination: in combination A all-site (1,436 evaluable blocks), 718 blocks showed positive local rho (mean +0.393) and 718 showed negative local rho (mean −0.396), with a median local rho of 0.0004 and 72 nominally significant windows (51–75 across combinations), excluding the possibility that a null global rg conceals offsetting positive and negative local signals (Supplementary Table S4).

### Bidirectional Mendelian randomization

Bidirectional two-sample MR across combinations A/B/C (knee, hip, all OA) used four estimators (IVW random-effects, weighted median, weighted mode, MR-Egger), four instrument settings (P<5×10⁻⁸ or P<1×10⁻⁶; clumping rZ<0.001 or <0.01), plus GSMR+HEIDI, MR-PRESSO, and TKR hard-outcome analyses (36 primary estimates: 3 combinations × 3 OA subtypes × 2 directions × IVW and weighted median; Fig. 4; Supplementary Tables S5–S6). Every OA → PD and PD → OA chain was null in the primary analysis (e.g., combination A all-site IVW β = −0.013, p = 0.92); PD → OA directions were nominally negative but never significant with consistent methods. The no-overlap combination A and the no-proxy combination E, the conditions under which the positive claim should be least biased, were also null. The 17q21 H1/H2 inversion sensitivity confirmed that MR estimates were not driven by MAPT-region instruments (OA → PD instrument sets contained no 17q21 instruments; PD → OA results were unchanged after removing rs199452). The OA × AD control showed similarly negative genetic correlations (rg −0.02 to −0.10 across primary and gcov-intercept-constrained estimates; e.g., gcov-intercept-corrected knee OA × AD rg = −0.077, p = 7.0×10⁻⁴), indicating the decoupling signature is not PD-specific (Supplementary Note N1).

**Fig. 4.**
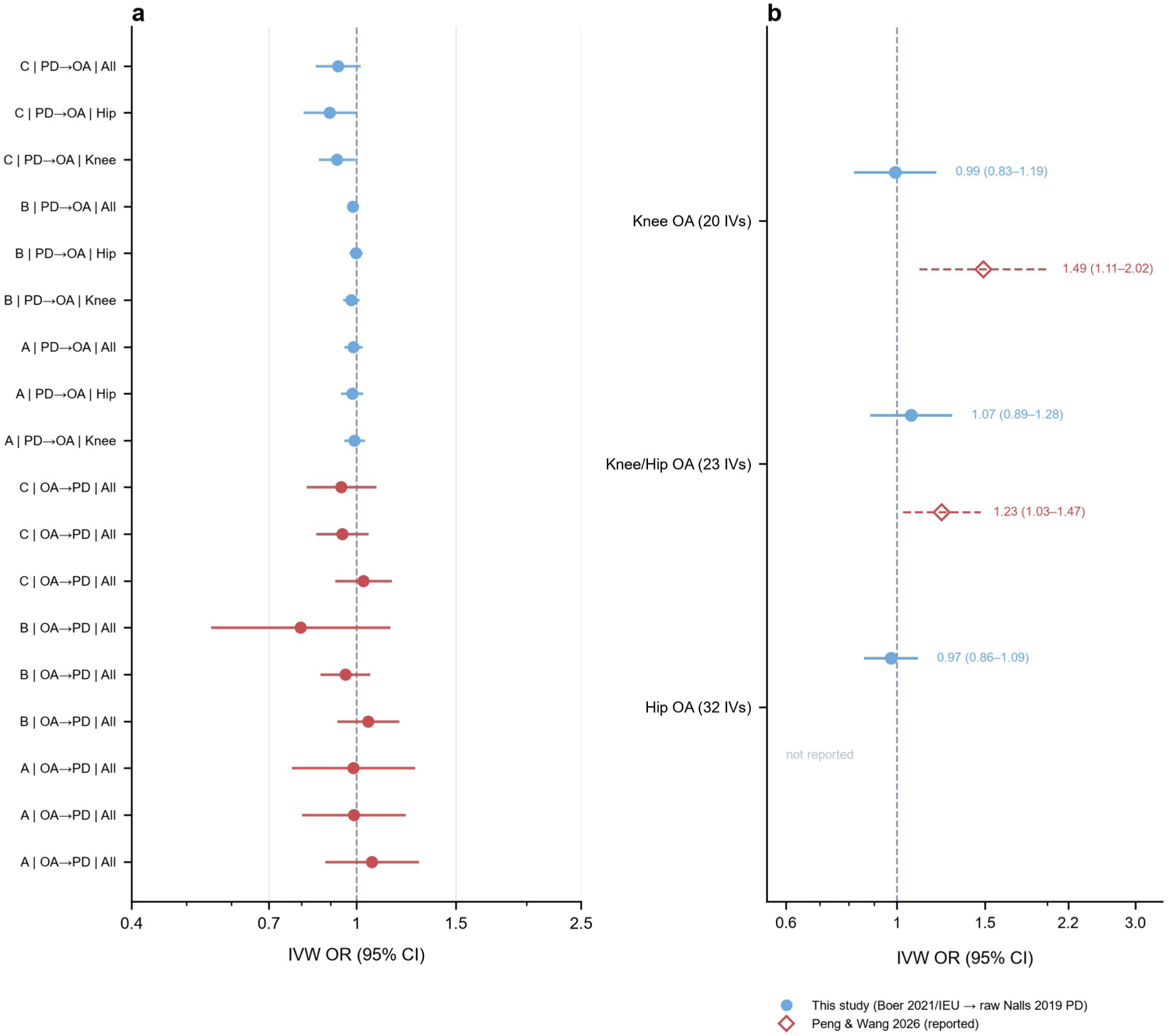
Bidirectional Mendelian randomization. (a) IVW forest plots for OA → PD (red) and PD → OA (blue) across combinations A/B/C × knee/hip/all (log scale; all 18 chains null); rows read “combination | direction | subtype”. (b) Combination D, instrument-set replication of Peng & Wang 2026 using the same underlying Boer 2021/IEU OA instruments against raw Nalls 2019 PD. Filled blue circles = this study (IVW random-effects): knee OA (20 IVs) OR 0.99 (95% CI 0.83–1.19); knee/hip OA (23 IVs) OR 1.07 (0.89–1.28); hip OA (32 IVs) OR 0.97 (0.86–1.09). Open red diamonds = Peng & Wang 2026 reported estimates: knee OA OR 1.49 (1.11–2.02); knee/hip OA OR 1.23 (1.03– 1.47); hip OA not reported. The two series are offset vertically within each row and every estimate is printed with its 95% CI, so overlapping CIs remain distinguishable. Although the knee-OA and knee/hip-OA CIs overlap numerically, all this-study point estimates are null and systematically lower, indicating that the positive MR was instrument-sensitive. Instrument sets were reconstructed from the Peng & Wang supplement and re-clumped with our QC pipeline (r²<0.001, 10 Mb, P<5×10⁻⁸, F>10, MHC excluded); they therefore share the same underlying databases but are not parameter-identical to the original analysis. Shared OA–PD instrument overlap across combinations is shown in Supplementary Fig. 6.

### Shared-locus mapping and colocalization

Conditional/conjunctional FDR (ccFDR) identified 732 independent shared loci at conjFDR<0.05 (A: 104; B: 64; E: 550; C: 14), with no directional bias in any combination (binomial p = 0.20–1.00; e.g., combination A all-site 21 same-vs 23 opposite-direction), indicating shared signals do not aggregate toward concordant or antagonistic effects (Fig. 5; Supplementary Tables S7–S8).

**Fig. 5.**
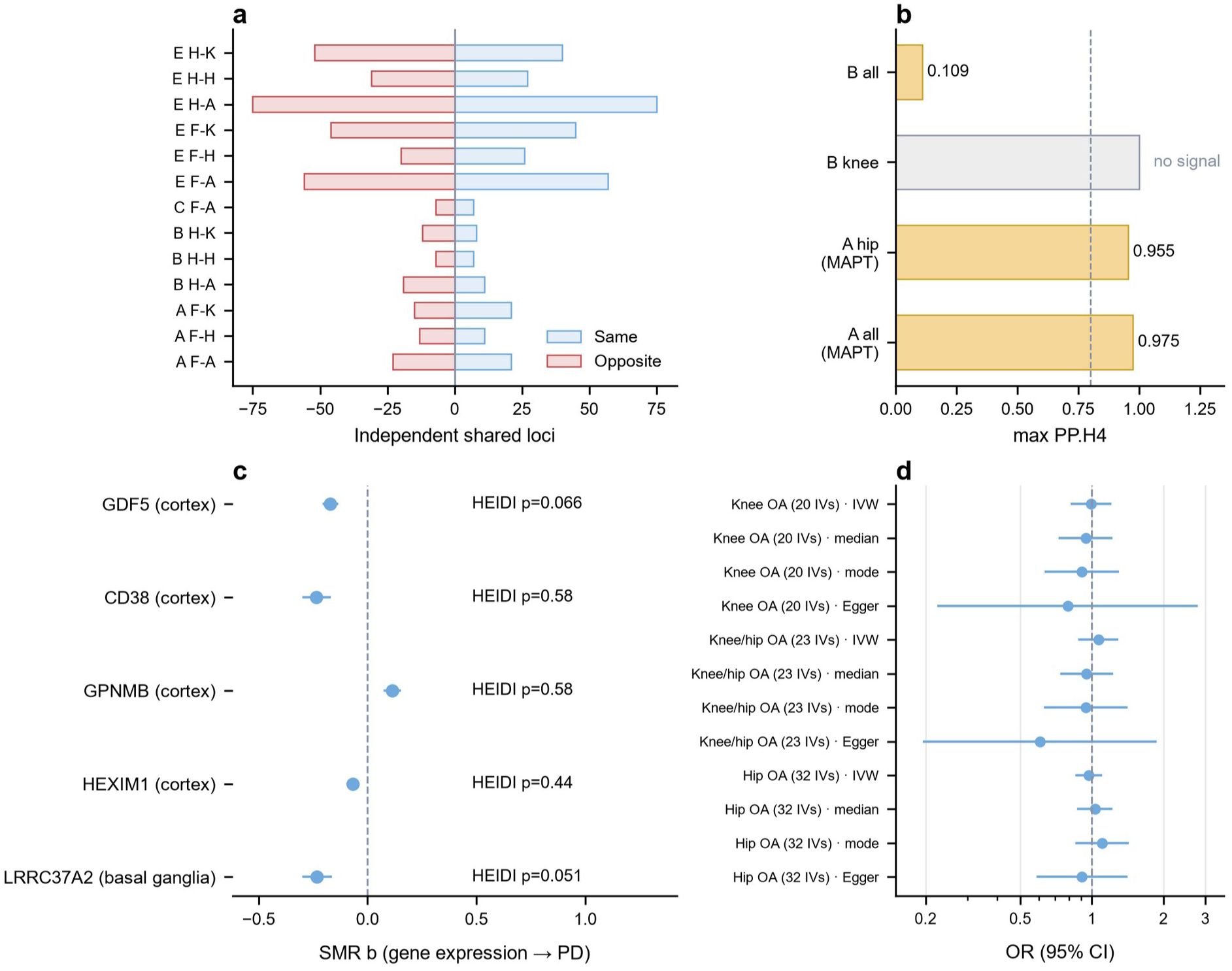
Shared loci and MAPT 17q21.31. (a) Direction of 732 ccFDR shared loci (conjFDR<0.05; A 104, B 64, C 14, E 550) as same-versus opposite-direction signals; rows read “combination | OA source–subtype” (F = FinnGen, H = Hatzikotoulas; K/H/A = knee/hip/all). No pair shows directional bias (binomial p = 0.20–1.00). D, F and G are absent because they were not in the ccFDR mapping: D is an instrument-set MR replication (20–32 variants), F a trans-ancestry PD update (EUR LD mismatch), G the OA × AD control. (b) SuSiE coloc maximum PP.H4 by pair (A all-site 0.975, A hip 0.955 at MAPT 17q21.31; B pairs no signal). (c) MAPT-region SMR effects (HEIDI-passing; HEXIM1 in MetaBrain cortex on the OA side, LRRC37A2 in basal ganglia on the PD side; positive controls GPNMB/CD38/GDF5; KANSL1 HEIDI-rejected). (d) Combination D MR across four estimators (IVW, weighted median, weighted mode, MR-Egger), all null (knee OA OR 0.99, 0.83–1.19; knee/hip 1.07, 0.89–1.28; hip 0.97, 0.86–1.09), confirming the OA → PD null is robust to instrument-set and estimator choice. ccFDR Manhattan: Supplementary Fig. 7.

SuSiE colocalization^17^ (allele-aligned) converged on a single robust region, MAPT 17q21.31 (maximum PP.H4 = 0.975 in combination A all-site; 0.955 in A hip); no other region met strong thresholds across combinations. SMR+HEIDI supported OA-side gene-level signals (HEXIM1 in MetaBrain^24^ cortex p = 1.8×10⁻⁶, HEIDI p = 0.435), with positive controls (GPNMB, CD38, GDF5) validating the pipeline; on the PD side, LRRC37A2 was SMR-significant (basal ganglia p = 2.7×10⁻¹¹, HEIDI p = 0.051), whereas the OA-side LRRC37A2 signal and KANSL1 SMR were HEIDI-rejected (LD-driven; p = 5.3×10⁻⁵ and LD-driven), consistent with PP3-dominant colocalization on the PD side. All MAPT-region evidence carries a haplotype warning (H1/H2 inversion); four inversion tags plus 2,960 LD proxies were annotated, and causal inference was unchanged after removing inversion-linked instruments (Supplementary Note N2).

The genome-wide architecture is therefore decoupled. The only robust shared focus, at MAPT 17q21.31, shows limited, direction-unbiased sharing (Fig. 5).

### Genetic mediation

Two-step multivariate MR^25^ across 24 chains (4 OA exposures × 6 mediator definitions/versions) found no significant indirect effects; step-2 conditional F statistics were 1.3–3.6 (all <10), pre-specifying “evidence insufficient (power-limited)” rather than “no effect.” Step-1 OA → chronic-pain instruments were adequate (conditional F 11.8–30.9). The genetic layer therefore cannot confirm the phenotypic depression mediation, which is consistent with these pathways being behavioural rather than strongly heritable (Table 4; Supplementary Table S10).

**Table 4.** Two-step multivariate MR (MVMR) genetic mediation, 24 chains.

| Layer | Result |
| --- | --- |
| Indirect effects (24 chains) | No significant indirect effect in any of the 24 pre-specified OA → mediator → PD chains |
| Step-2 conditional F statistics | 1.3–3.6 (all <10; weak instruments) |
| Step-1 OA → chronic-pain instruments | Conditional F 11.8–30.9 (adequate) |
| Pre-specified conclusion | Evidence insufficient (power-limited), not evidence of absence |

### Cell-type-specific layer

Cell-type-specific MR (csMR; Bryois 2022 eight brain cell types) identified 71/42 (hits/genes) FDR-significant gene–PD associations with Nalls 2019^20^, 104/73 with IPDGC no-UKBB^22^, 47/33 with FinnGen PD, and 68/48 with Kim 2024^23^; MAPT-region genes (KANSL1, LRRC37A2, ARL17B, CRHR1) showed consistent cross-cell negative effects, matching the PD-side cell-type MR direction^13^. However, MR-Egger intercepts were significant in 18/68 evaluable candidate analyses (72 candidate rows; Supplementary Table S11), indicating pervasive horizontal pleiotropy; three-trait colocalization (OA × eQTL × PD) found only 7 rows with bilateral PP.H4>0.8 (4 gene–cell pairs: KANSL1 in endothelial cells and oligodendrocytes; TTLL4 and CYP27A1 in microglia), and the closed chain in the no-overlap combination A was zero under the pre-specified criteria (formal multi-instrument candidate, bilateral PP.H4>0.8, direction consistency): endothelial KANSL1 was PP.H3-dominant (PP.H4 = 0.12) on the PD side, oligodendrocyte KANSL1 showed bilateral colocalization but was not a formal multi-instrument candidate, and all 17q21 signals carry the H1/H2 haplotype warning. Single-cell disease-relevance scoring (scDRS) enriched OA signals in astrocytes and PD signals in microglia (FDR≈0; PD also astrocyte-enriched, FDR = 6.7×10⁻¹¹⁵) but not in dopaminergic neurons; EWCE gave only nominal support (Supplementary Fig. 4).

Cell-type-specific evidence thus confirms MAPT-region involvement across brain cell types but does not close the “OA risk locus → brain-cell expression → PD” chain; the csMR signal is more consistent with LD/pleiotropy (Fig. 6; Supplementary Table S11).

**Fig. 6.**
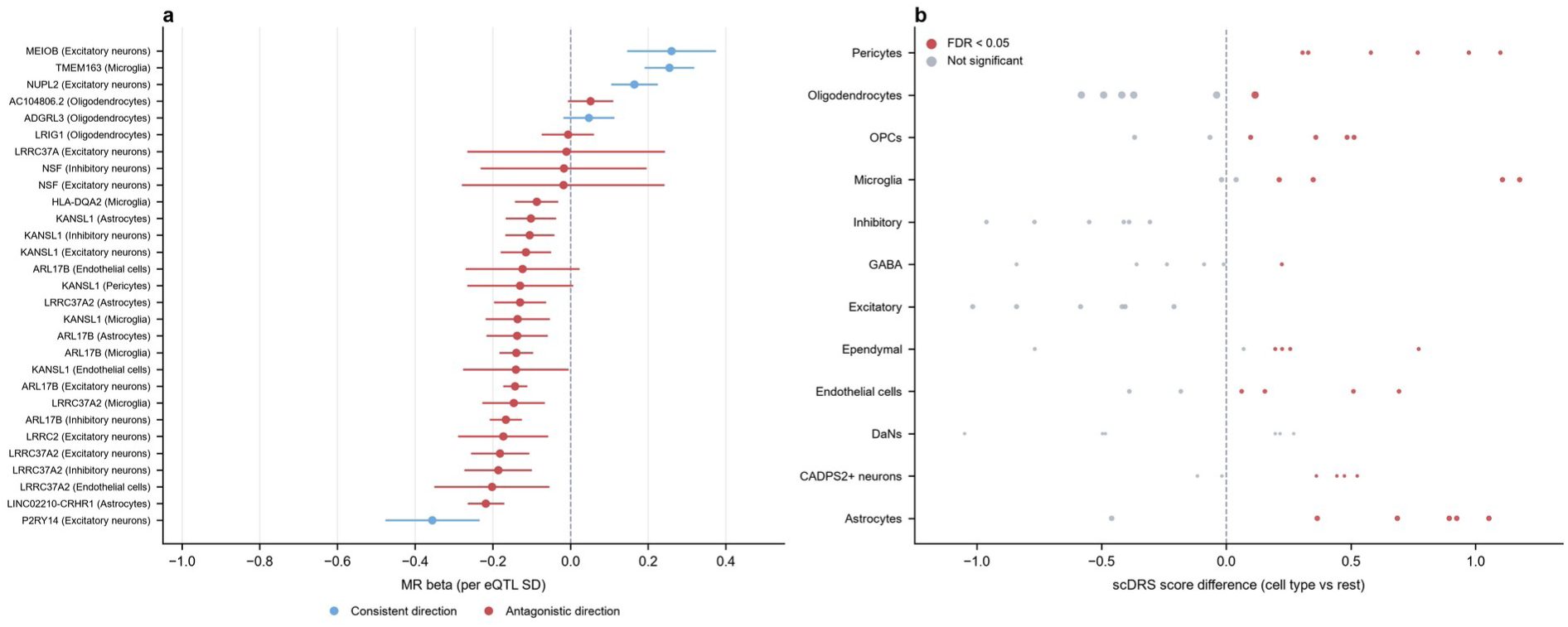
Cell-type-specific layer. (a) csMR gene–PD forest plots for the Nalls 2019 outcome (all 29 FDR<0.05 strict multi-instrument candidates; blue = consistent, red = antagonistic direction). (b) scDRS cell-type enrichment in PD midbrain snRNA-seq (red = FDR<0.05; OA enriched in astrocytes, PD in microglia). Three-way direction heatmap and MAGMA gene-level results are shown in Supplementary Figs. 8–9; csMR sensitivity estimates across the four PD GWAS are shown in Supplementary Fig. 5.

### Expression validation

Direction consistency between cell-type-specific genetic predictions and differential expression was 63.8% (51/80) in Smajić 2022, 75% (42/56) in Martirosyan 2024, and 30.6% (45/147) in Kamath 2022 (coarse annotation, limited genetic-direction coverage). Single-cell MAST identified 1 significant pair in Smajić and 7 in Martirosyan (e.g., LRRC37A2 in microglia, NSF in astrocytes/neurons), with no consistent cell-type pattern. These results do not support a strong expression-mediated pathway (Fig. 7; Table 5; Supplementary Table S12).

**Fig. 7.**
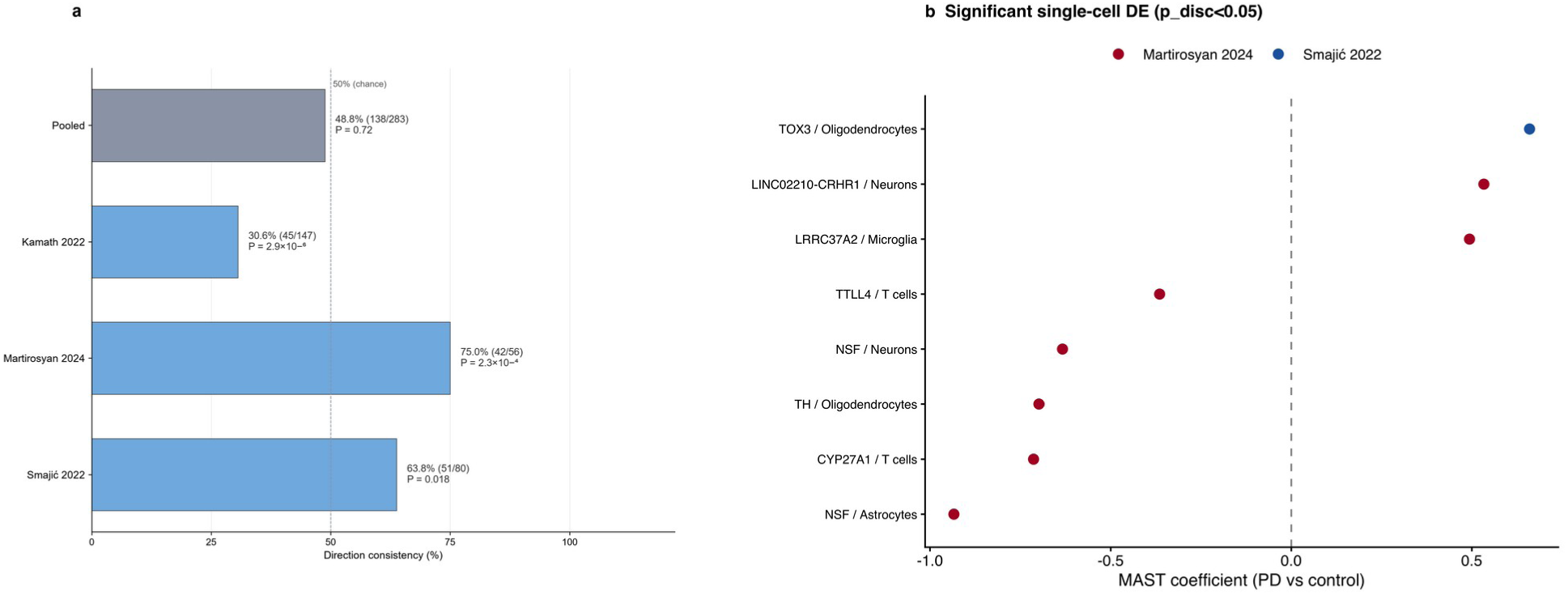
Expression validation. (a) Direction-consistency rates with two-sided exact binomial tests against 50% (Smajić 63.8% [51/80], p = 0.018; Martirosyan 75.0% [42/56], p = 2.3×10⁻⁴; Kamath 30.6% [45/147], p = 2.9×10⁻⁶; pooled 48.8% [138/283], p = 0.72); the opposing directions across datasets indicate no reproducible direction signal. (b) Significant single-cell MAST differential expression (p_disc<0.05, PD vs control), coefficient with 95% CIs. Pseudobulk DESeq2 results are shown in Supplementary Fig. 10.

**Table 5.** Direction consistency between genetic predictions and differential expression.

| Dataset | Genes with expected direction | Direction-consistent | Consistency |
| --- | --- | --- | --- |
| Smajić 2022 | 80 | 51 | 63.8% |
| Martirosyan 2024 | 56 | 42 | 75.0% |
| Kamath 2022 | 147 | 45 | 30.6% |

### Functional enrichment

Functional enrichment analyses (exploratory; documented amendment, Supplementary Note N4) further distinguished the OA and PD signals. MAGMA gene-set analysis of the MSigDB v7.4 GO/KEGG collections identified strong OA-specific enrichment in cartilage biology: chondrocyte differentiation (BH-FDR = 6.4×10⁻⁸ for Hatzikotoulas all OA and 1.7×10⁻⁵ for FinnGen knee OA) and cartilage development were the leading terms, with 2–50 pathways surviving BH-FDR across the four OA traits; neither PD GWAS yielded an FDR-significant pathway (smallest gene-set p = 1.7×10⁻⁵, IPDGC no-UKBB). DGIdb drug-target categories showed the same contrast: OA signals were enriched in tyrosine kinases (FDR = 9.9×10⁻3 for FinnGen all OA), transcription-factor complexes, clinically actionable genes and approved-drug targets (1–4 categories with FDR<0.05 per OA trait), whereas PD showed none. GTEx v8 tissue-expression analysis (54 tissues) found a broad positive correlation with OA signal: 52/54 tissues passed BH-FDR for knee OA and 38–42 tissues for the all-OA traits, with brain regions strongest (cerebellum p = 2.1×10⁻⁸); PD showed no significant tissue correlation. The 141 csMR candidate genes (including the cross-cell-consistent 17q21 genes) were not enriched in any GO/KEGG term or DGIdb category after correction, indicating that the shared signal lacks a broad pathway, drug-target, or tissue-specific signature (Fig. 8; Supplementary Tables S13–S15).

**Fig. 8.**
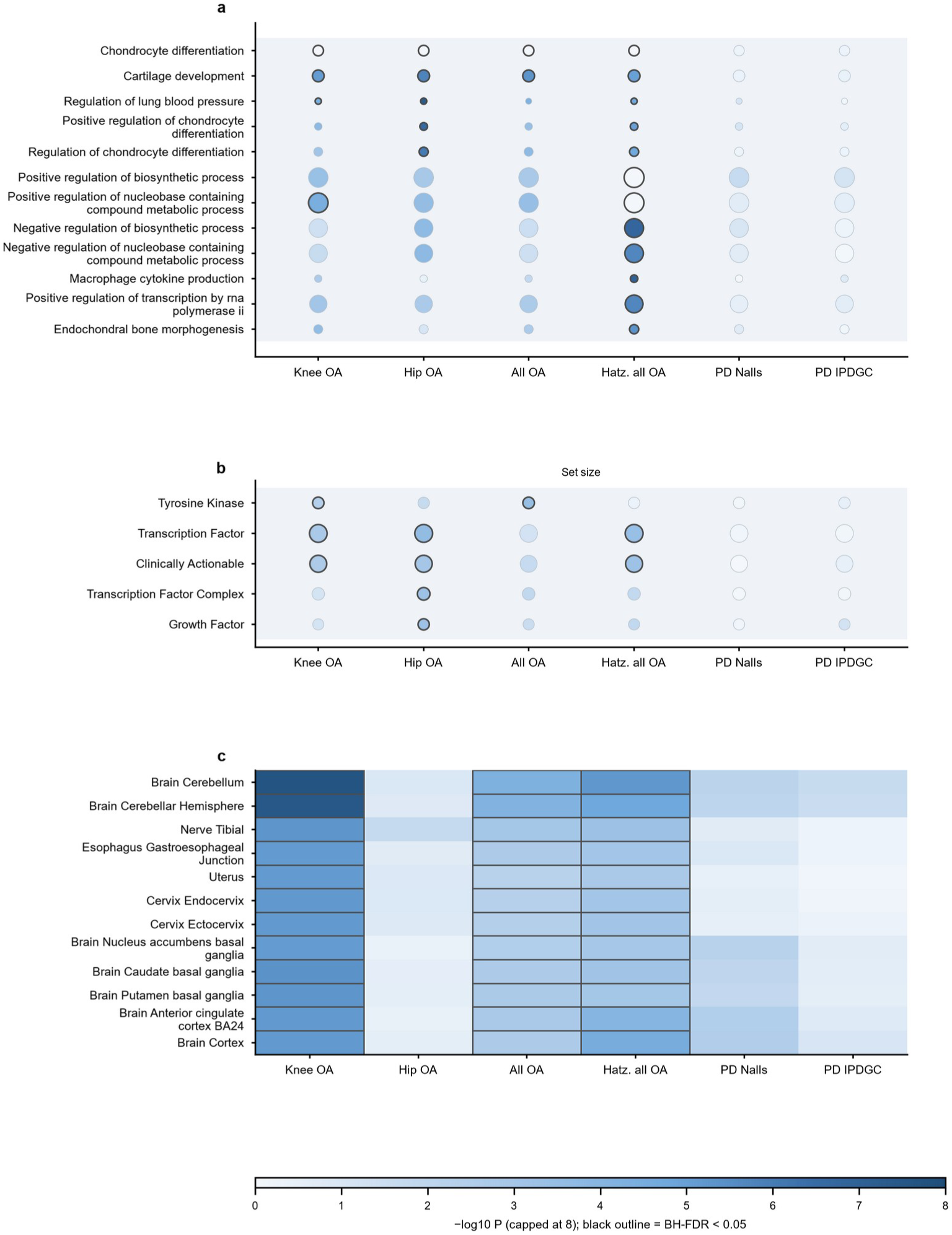
Functional enrichment of OA and PD GWAS signals. (a) MAGMA competitive gene-set enrichment of MSigDB v7.4 GO/KEGG terms (top 12 sets with BH-FDR<0.05 in ≥1 trait, rows hierarchically clustered; dot area ∝ set size; colour = −log10 P; light-grey bands denote GO domain). (b) MAGMA enrichment of DGIdb v5.0.11 drug-target categories (top 5 FDR-significant categories, same encoding). (c) MAGMA GTEx v8 tissue-expression enrichment (top 12 of 54 tissues, rows clustered within tissue groups; colour = −log10 P capped at 8; outlined cells = BH-FDR<0.05). Columns: FinnGen knee/hip/all OA, Hatzikotoulas all OA, Nalls 2019 PD, IPDGC no-UKBB PD. OA signals are enriched in cartilage-development pathways, druggable categories and broad tissue expression; PD shows no significant enrichment in any layer; csMR candidate genes show no pathway or drug-target enrichment (Supplementary Tables S13–S15).

These annotations reinforce the genetic-decoupling conclusion: OA risk loci are enriched in cartilage and druggable biology and correlate with expression across most tissues, whereas PD loci show no such enrichment and the only robust shared focus (17q21) carries no pathway, drug-target, or tissue-specific signature. Along with the cell-type and expression layers, they provide no support for an expression-or pathway-mediated OA → PD mechanism beyond the MAPT 17q21.31 region.

## Discussion

This study provides, to our knowledge, the first six-layer triangulation of the OA–PD association. The results converge: the observational association is reproducible but non-specific, proxy-sensitive, and substantially depression-mediated (consistent with a phenotypic illusion), whereas the genetic architecture is decoupled genome-wide, with only limited, direction-unbiased sharing at MAPT 17q21.31 and no support for an expression-mediated causal pathway.

Our findings directly address the positive MR claim by Peng and Wang^10^ (knee OA OR 1.49). That analysis, like earlier positive reports^9^, used a limited instrument set without systematic sample-overlap auditing or no-proxy-PD sensitivity. In our data the association disappears under conditions removing the two most plausible biases: complete sample independence (combination A, FinnGen OA × Nalls 2019 PD^20^) and removal of proxy cases (combination E, IPDGC no-UKBB^22^). A direct instrument-set replication (combination D), rebuilt from the same underlying Boer 2021/IEU^26^ OA instruments, was also null (knee OA IVW OR 0.99, 95% CI 0.83–1.19; Fig. 4b), and our genetic-correlation estimates (null to negative) are inconsistent with a causal OA → PD effect. We emphasise that the positive finding is not robust to stricter design choices rather than that it is “wrong.”

Wang et al.^11^ combined survival analysis, MR, transcriptomic mapping, and in vivo perturbation to study the arthritis–neurodegeneration axis. They reported OA-associated risks of AD, PD, and autonomic-nervous-system disease and an RA-associated AD risk; observationally, RA was not associated with PD, although MR suggested a modest protective RA → PD effect (OR 0.93, 0.88–0.99), and RNF40 function was context-dependent. Our study adds the OA-exposure-side genetic-structure evidence (LDSC, LAVA, ccFDR, colocalization, MVMR) that Wang et al. did not report; both studies agree that OA is not a strong causal driver of PD risk.

Our cell-type-specific results intersect with the PD-side cell-type MR by Gu et al.^13^ (ARL17A/ARL17B, KANSL1, LRRC37A across brain cell types): we observe the same MAPT-region genes with consistent cross-cell directions from the OA exposure side. However, three-trait colocalization did not close the chain from OA loci through brain-cell expression to PD, and MR-Egger indicated pervasive horizontal pleiotropy. The PD-side signal is therefore best interpreted as a property of the MAPT 17q21 region relevant to PD itself, not as OA risk alleles acting through those cells to cause PD.

The MAPT 17q21.31 region is a large H1/H2 inversion with pleiotropic associations across multiple neurodegenerative diseases. Our robust colocalization (PP.H4 = 0.975) with direction-unbiased shared loci, together with SMR/HEIDI support for HEXIM1 on the OA side and LRRC37A2 on the PD side, establishes a focused, testable hypothesis for the OA–PD interface (e.g., inflammation, autophagy-lysosomal, or stress-response biology). The absence of directional bias and the failure of expression-level validation caution against over-interpreting the region as an OA-specific causal mediator; we therefore describe it as limited, direction-unbiased sharing rather than antagonistic or protective. A recent npj Parkinson’s Disease triangulation of hearing loss and PD reported localized genetic overlap at 17q21.31 ^27^, supporting this region as a general pleiotropic hub for age-related conditions rather than an OA-specific interface.

From a clinical and public-health perspective, our results argue against the premise that joint-directed OA treatment would meaningfully reduce PD risk. Instead, the consistent depression-mediated component across two cohorts supports depression screening and comorbidity management in patients with OA as the more actionable preventive strategy. These recommendations are hypothesis-generating: the genetic-layer mediation was power-limited (weak instruments), and the phenotypic mediation rests on observational designs.

Strengths include triangulation across evidence layers with non-overlapping bias structures; a complete sample-overlap audit (no-overlap A, no-proxy E, gcov-intercept sensitivity); allele-aligned SuSiE colocalization^17^ with standard conditional FDR; MAPT H1/H2 inversion sensitivity; an OA × AD contrast; and version-pinned analyses with the full protocol archived in OSF.

This study has several limitations. First, NHANES contained only 20 strict PD-proxy events, limiting precision and specificity inference. Second, the two cohorts used different outcome definitions and were not pooled. Third, MVMR was underpowered, pre-specifying “evidence insufficient” rather than “evidence of absence.” Fourth, the Bryois eQTL data lack midbrain dopaminergic neurons, providing cortical neuronal evidence only. Fifth, Kamath 2022 annotation was coarse, with low direction consistency. Sixth, the Kim 2024^23^ PD GWAS is trans-ancestry, with imperfect EUR LD reference matching. Seventh, the CAUSE^28^ elpd comparison layer could not be completed under the installed R environment; horizontal pleiotropy was instead covered by MR-Egger, MR-PRESSO, and GSMR+HEIDI (Supplementary Note N3). Eighth, two-sample MR assumes a linear exposure–outcome relation; nonlinear MR requires individual-level data and was not possible with the summary-statistics framework used here. The phenotypic layer rests primarily on CHARLS: only CHARLS reached significance for the total association, whereas NHANES contributed a directionally consistent total effect and significant, directionally consistent mediation support. Under the decision rule specified in the analysis protocol, a single-cohort signal is downgraded to exploratory unless the second cohort provides such support (documented amendment; Supplementary Note N4); accordingly, the phenotypic layer is interpreted as supportive rather than confirmatory. The instrument-set replication of Peng & Wang (combination D) used OA instruments reconstructed from the official supplement because the original paper’s full methods and supplementary materials are not openly available, so exact parameter-for-parameter identity cannot be guaranteed.

The OA–PD association observed in epidemiological studies is largely consistent with a phenotypic illusion: reproducible, non-specific, proxy-sensitive, and substantially depression-mediated, with genome-wide genetic decoupling and only locus-restricted, direction-unbiased sharing at MAPT 17q21.31. Future work should test the MAPT-region hypothesis in dopaminergic-neuron eQTLs and functional models, extend the decoupling contrast across the OA–AD–PD matrix, and evaluate depression and comorbidity management in OA as candidate preventive targets.

## Methods

### Study design, registration, and reporting

This is a six-layer triangulation study of the OA–PD association, organised in the Methods into four evidence blocks: (i) a phenotypic layer using two population cohorts (CHARLS and NHANES); (ii) a genome-wide genetic architecture layer (genetic correlation, bidirectional Mendelian randomization [MR], shared-locus mapping, and multivariate MR [MVMR]); (iii) a cell-type-specific layer (cell-type-specific MR [csMR] and single-cell enrichment); and (iv) a single-cell expression validation layer using three PD midbrain single-nucleus RNA-sequencing (snRNA-seq) datasets. The analytical workflow is summarised in Supplementary Fig. 1. The study design, variable definitions, models, thresholds, and decision rules were developed and archived in the OSF project (https://osf.io/rgeaf; registration DOI 10.17605/OSF.IO/K8Q5A) before the analyses were finalized. Because analyses had begun before registration, the OSF registration (completed 25 August 2026) is retrospective rather than prospective. Methodological amendments, including the use of standard conditional FDR, allele-aligned SuSiE colocalization^17^, pinned random-effects IVW versions, candidate gene re-screening, MAPT 17q21 haplotype annotation, Bryois brain-region wording, and downgrading of Kamath annotation, are documented in Supplementary Note N4. The phenotypic layer is reported in accordance with the STROBE statement and the genetic layers in accordance with the STROBE-MR checklist^29^. All analyses used publicly available, de-identified secondary data; no new data collection or ethics approval was required (confirmed in writing per institutional policy).

### Phenotypic data sources and variables

#### CHARLS (2011–2020)

We used the China Health and Retirement Longitudinal Study (CHARLS)^30^, waves 2011 (baseline), 2013, 2015, 2018, and 2020, restricted to participants aged ≥50 years at baseline with no baseline stroke (da007_8_=1) or memory-related disease (da007_12_=1). The KOA proxy was defined as self-reported arthritis/rheumatism in 2011 (da007_13_=1) combined with knee pain at site code 12 (da042s12==12), and was reviewed using the 2020 arthritis item (da003_14_). The primary PD outcome was the independent 2020 chronic-disease item for Parkinson’s disease (da003_13_=1); new-onset PD required da002_13_==99 (“never had” relative to ZIWTime). Alternative outcomes were memory-related disease (2013–2018: da007_12_ including PD; 2020: da003_12_ excluding PD) and informant-reported PD in 2018 (dd009_w4, a subpopulation of proxy interviews).

Mediators were depression (CES-D-10 ≥10, dc009–dc018 with reverse scoring for dc013/dc016) and a sleep score derived from sleep duration (da049) and restlessness (dc015) [exploratory]. Covariates were age, sex, education (bd001, four categories), marriage (be001∈{1,6}), log per-capita income (INCOME_PC), measured BMI (2011), smoking (da059=1), alcohol use (da067=1), and comorbidity count. Standard errors were clustered by community (communityID). CHARLS cannot distinguish rheumatoid arthritis (recorded as a limitation).

#### NHANES (2011–2018)

We used four cycles (2011–2012, 2013–2014, 2015–2016, 2017–2018; G–J) of the National Health and Nutrition Examination Survey (NHANES)^31^, a cross-sectional survey. Participants were restricted to those aged ≥50 years with a non-missing MEC examination weight >0. Knee OA (KOA) was defined as self-reported physician-diagnosed arthritis (MCQ160a=1) with a reported type of osteoarthritis (MCQ195=1); the control group comprised participants reporting no arthritis (MCQ160a=0). Specificity analyses used four mutually exclusive exposure categories: KOA, rheumatoid arthritis (MCQ195=2), other arthritis (MCQ195∈{3,4}), and no arthritis.

The primary PD-specific outcome was defined by prescription anti-Parkinson drugs in the prescription medication files (RXQ_RX): levodopa, MAO-B inhibitors, or COMT inhibitors (combination products counted if containing levodopa), excluding participants using amantadine or restless legs syndrome (RLS) agonists (primary analytic sample n = 6,320, events = 20); a levodopa-only definition (n = 6,320, events = 18) was used as an additional stricter sensitivity. Proxy-control outcomes included RLS drugs (pramipexole, ropinirole, rotigotine) and anti-dementia drugs (donepezil, rivastigmine, galantamine, memantine), which are known to cause misclassification if mixed into a PD proxy. Negative-control outcomes were self-reported asthma (MCQ010) and hearing difficulty (AUQ054≥3, available in cycles G/I/J).

Mediators were depression (PHQ-9 total score ≥10, based on DPQ010–DPQ090 with ≥7 valid items), physical inactivity (answering “no” to all PAQ605/620/650/665 items), and short sleep (<7 h; SLD010H in cycles G/H, SLD012 in I/J); extended mediation analyses additionally included obesity (BMI ≥30) and cardiovascular comorbidity. Covariates were age, sex, BMI ≥30 kg/mZ, White race, smoking (≥100 lifetime cigarettes), alcohol use (≥1 drink/month in the past 12 months), comorbidity count (congestive heart failure, coronary heart disease, angina, myocardial infarction, stroke), and diabetes (self-reported diagnosis or glycemic criteria). Four-cycle weights (WTMEC2YR/4) were used, with dual-track sensitivity using interview weights (WTINT2YR).

### Phenotypic analyses

The two cohorts were analysed in parallel without pooling raw data, using the same directed acyclic graph (DAG), the same model structure (M1–M4), and the same output table structure. Mediators (depression, sleep, and physical inactivity in NHANES, with obesity and cardiovascular comorbidity in extended analyses; depression and incident stroke in CHARLS multi-mediator decompositions) were treated as mediators rather than confounders: the primary models estimated the total effect without adjustment for mediators, and mediation was quantified separately. Age, sex, BMI, smoking, and comorbidities were treated as confounders and adjusted.

CHARLS models. Logistic regression with cluster-robust standard errors (communityID; sandwich/clubSandwich) was the primary model, with the same M1–M4 structure (M4 adding comorbidity count); Firth penalization was used as a small-sample check.

NHANES models. Survey-weighted logistic regression was the primary inference model, with Firth penalization as a small-sample sensitivity to accommodate sparse events:

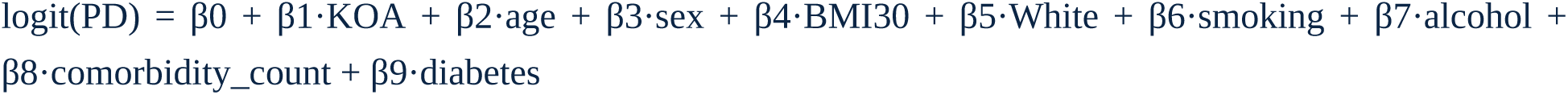

M1 crude; M2 +age/sex/BMI30/White; M3 +smoking/alcohol; M4 +comorbidity count/diabetes. Sensitivity analyses used survey-weighted logistic regression (survey svglm) with PSU/strata/weights; PSU-bootstrap confidence intervals were archived as exploratory only, because sparse events can degrade CI coverage.

Phenotypic-illusion tests (H1). (i) Attenuation test: mediators were added one at a time to M4, and the percentage attenuation of the OR was compared, with a pre-specified 15–20% attenuation considered “marked.” (ii) Formal causal mediation: the mediation R package with 300 bootstrap iterations (200–300 for NHANES) was used, reporting NDE/NIE/proportion-mediated with CIs; NHANES added survey-weighted sensitivity. For CHARLS, a temporal design was used (2011 baseline KOA exposure, 2013/2015 depression mediator, 2020 PD outcome) so that the mediator strictly preceded the outcome; both the “add mediator to M4” attenuation and the formal NDE/NIE proportion are reported. (iii) Specificity: the four exposure categories were entered simultaneously (NHANES), with Fisher’s exact OR as a small-sample check; negative outcomes (asthma, hearing) and proxy-control outcomes (RLS drugs, anti-dementia drugs) were analysed. (iv) E-values were computed for unmeasured confounding (VanderWeele & Ding 2017^18^). (v) Stratified analyses by age (<65/≥65), sex, and NHANES cycle; strata with <10 events are reported for direction only.

Competing risk and attrition (CHARLS). Death and attrition were reconstructed from wave exits and informant modules; the primary analysis was followed by a Fine-Gray subdistribution hazard model (death as competing event) and inverse-probability weighting (IPW), with attrition bounds reported.

Decision rules (pre-specified in the analysis protocol). The “phenotypic illusion” conclusion is supported when the OA total effect is significant, adjustment for mediators markedly attenuates the effect, and non-OA arthritis/negative outcomes show similar associations, with direction consistency across cohorts and a pre-specified mediation proportion ≥30% (when estimable). If only one cohort reaches significance for the total effect, the finding is downgraded to exploratory unless the second cohort provides (i) a directionally consistent total effect and (ii) a significant, directionally consistent mediation effect, in which case the layer is retained as supportive but described with calibrated language (documented amendment; Supplementary Note N4).

### Genetic data sources, combinations, and quality control

Data matrix (combinations A–G, frozen in Protocol v4.4). A = FinnGen R13^32^ OA (knee 64,535, hip 32,962, all 106,674 cases) × Nalls 2019 PD^20^ (non-23andMe; GCST009325) [level I, no overlap; primary causal evidence]. B = Hatzikotoulas 2025 EUR OA^21^ (knee 161,228, hip 95,395, all 441,288 cases; GCST90566795/798/800) × Nalls 2019^20^ [level II, UK Biobank overlap quantified by gcov; maximum power]. C = FinnGen R13^32^ OA × FinnGen R13^32^ G6_PARKINSON (6,320 cases/493,866 controls) [level III, same-population consistency]. D = Boer 2021/IEU^26^ OA instrument sets × Nalls 2019^20^ [confrontation of published positive MR; all null, full results in Methods]. E = OA (FinnGen or Hatzikotoulas) × IPDGC 2021 no-UKBB^22^ PD (males 12,054/11,999, females 7,384/12,389; meta ≈19,438 cases/24,388 controls; kp4cd nodes 1291/1290) [no-proxy sensitivity, near-complete independence]. F = OA × Kim 2024 GP2^23^ PD, trans-ancestry, without 23andMe (44,358 cases + 18,618 proxy/966,017 controls; kp4cd node 1127) [updated PD version; EUR LD mismatch acknowledged, conservative interpretation]. G = OA × Bellenguez 2022 AD^33^ (GCST90027158; 111,326 cases + 67,703 proxy/677,874 controls) [disease-control contrast]. Mediator GWAS: chronic pain Johnston 2019^34^ (GCST008512), PGC MDD 2023+ (mdd2023diverse^35^), insomnia Watanabe 2022^36^ (kp4cd Watanabe2022_Insomnia_EU; GCST90131901 unavailable on EBI FTP), physical activity Klimentidis 2018^37^ (GCST006097), BMI Yengo 2018^38^ (ieu-b-40).

All OA sources are nested (not independent replicates); per the frozen protocol, no fixed-effect meta-analysis across OA sources was performed, and evidence is presented as single-combination estimates plus an overlap matrix and direction-consistency summary. The UK Biobank overlap between Hatzikotoulas OA and Nalls 2019 PD^20^ (proxy cases account for 55% of Nalls cases, 18,618/33,674) is disclosed and quantified via the LDSC gcov intercept.

Quality control pipeline. All summary statistics were harmonized to SNP A1 A2 BETA SE Z P N with uppercase alleles; non-ACGT variants and duplicate SNPs were removed; strand-ambiguous (A/T, C/G) variants without unambiguous allele-frequency information were excluded during MR harmonization, and palindromic variants with unambiguous allele frequencies were resolved using effect-allele frequencies. GRCh37 data were used directly; GRCh38 data (FinnGen) were matched by rsID and LD reference. Beta values were used preferentially; log(OR) was used when only ORs were provided; when SE was missing, it was reconstructed as SE=|beta|/|z| from p (method recorded and validated for self-consistency; Hatzikotoulas official files lack an SE column and were processed this way). Official case/control counts were used (--N-cas/--N-con). All raw files were md5-verified with version IDs and download dates recorded in sources_manifest.csv. QC metrics (λGC, mean χZ, LDSC intercept, and LD-reference merge rate, expected >90%) were reported per file.

### Genetic architecture analyses

#### LD score regression (LDSC)

Single-trait heritability was estimated on the observed and liability scales (population prevalence: PD 1%, OA 10%) using LD score regression^19^ (LDSC 2.0.1) with 1000 Genomes Phase 3^39^ EUR LD scores (Zenodo 8182036). Pairwise genetic correlation (rg) was estimated for each OA subtype (knee/hip/all) within each combination. Sample overlap was handled by freely estimating the gcov intercept, with a sensitivity version constraining --intercept-gencov 0 for known-overlap combinations. Multiple testing was controlled by Bonferroni across combinations × OA subtypes × PD versions, with unadjusted values also reported; rg is reported with 95% CIs.

#### Local genetic correlation (LAVA)

Local genetic correlation was estimated with LAVA^40^ across 2,495 genomic blocks (a subset evaluable per combination because of SNP-coverage requirements; e.g., 1,436 in combination A all-site), to detect regions where local correlation is non-zero despite a null global rg. Local tests were corrected for the number of windows (FDR).

#### Bidirectional two-sample MR

Instrumental variables were genome-wide significant (P<5×10⁻⁸), independent (rZ<0.001, 10 Mb window; PLINK 1.9 clumping with 1000G EUR reference), with F>10, excluding the MHC region (chr6: 25–34 Mb). The primary method was inverse-variance weighted (IVW; random effects), with weighted median (bootstrap SE), weighted mode, and MR-Egger as sensitivity estimators. Robustness analyses included Cochran’s Q (heterogeneity), MR-Egger intercept (horizontal pleiotropy), leave-one-out, MR-PRESSO (outliers), Steiger directionality, and MR-RAPS. In addition, GSMR with HEIDI filtering (LD-clustered instruments with pleiotropy filtering) was run. CAUSE^28^ was attempted for joint modelling of correlated and uncorrelated horizontal pleiotropy; the mixSQP parameter estimation succeeded (74,879 LD-pruned SNPs), but the elpd causal-comparison layer failed because of incompatibility between the cause/loo stack and R 4.6; this is formally declared, and horizontal pleiotropy is covered by MR-Egger, MR-PRESSO, and GSMR+HEIDI (optional CAUSE^28^ rerun under R≤4.4 is documented).

Combination priority: A (no overlap) = primary causal evidence; B (maximum power, UK Biobank overlap disclosed) = main estimate; C = internal consistency. Reverse MR (PD → OA) used the identical pipeline.

Confrontation experiment (combination D). To test whether the published positive knee-OA-to-PD MR (Peng & Wang 2026^10^) is sensitive to instrument selection, we reconstructed the same underlying Boer 2021^26^ OA instrument sets from the paper’s official supplementary tables (Table S3 in Peng & Wang: 100 genome-wide-significant signals; Table S5: allele annotations; alleles were recovered across phenotypes by beta-direction consistency) because the IEU OpenGWAS VCF files (ieu-b-27/28/29) were unavailable (API retired/404). Instruments were clumped at rZ<0.001 within 10 Mb against 1000 Genomes Phase 3 EUR, restricted to P<5×10⁻⁸ and F>10, and removed from the MHC region, using the same pipeline as the primary analyses. The outcome was the full raw Nalls 2019 PD^20^ summary statistics (GCST009325; 33,674 cases/449,056 controls, excluding 23andMe), with 267 of 279 requested variant positions recovered (the LDSC-munged HapMap3 subset used elsewhere covers only 4 of the 22 original knee instruments; 20 passed our QC). MR was performed with IVW (random effects), weighted median, weighted mode, and MR-Egger, with Cochran Q, Egger-intercept, and MR-PRESSO sensitivity analyses. Knee OA (20 instruments): IVW OR 0.99 (95% CI 0.83–1.19, p = 0.93); knee/hip OA (23 instruments): OR 1.07 (0.89–1.28, p = 0.49); hip OA (32 instruments): OR 0.97 (0.86–1.09, p = 0.64); all sensitivity estimators were null, with no significant heterogeneity or pleiotropy and no MR-PRESSO outliers (full results in Supplementary Table S5; Figs. 4b and 5d).

#### Conditional/conjunctional FDR (ccFDR)

Conditional FDR and conjunctional FDR were computed bidirectionally (Andreassen 2013^41^; Smeland 2020^42^) with the standard conditional-FDR estimator (empirical conditional CDF via a Fenwick tree; Storey π0), implemented in Python (see Supplementary Note N4). Shared loci were defined at conjFDR<0.05 (primary) and <0.01 (strict) and deduplicated at rZ<0.1. Directional analysis compared same-direction versus opposite-direction effects using a two-sided exact binomial test. Output lists (SNP, direction, both P values) were the input for colocalization and downstream cell-type-specific analyses.

#### Colocalization (SuSiE-coloc)

Colocalization was performed in regions defined as ccFDR shared loci ±1 Mb. SuSiE-coloc ^17^ (coloc 5.2.3 + susieR) was the only primary colocalization method; coloc.abf produced NaN under the installed environment and was not used for inference. PP.H4>0.8 defined strong colocalization and 0.5–0.8 suggestive throughout; PP.H4>0.75 was used only as a screening threshold for candidate regions, with final inference based on the primary thresholds (PP.H4>0.8 strong / 0.5–0.8 suggestive), with H3/H4 probabilities reported and multi-signal regions decomposed by SuSiE.

#### MAPT 17q21.31 inversion sensitivity

The MAPT region (17q21.31) lies in a large H1/H2 inversion; all shared/colocalization evidence in this region carries a haplotype warning. Four inversion tags (rs8070723, rs17563986, rs393152, rs9468) plus 2,960 LD proxies (rZ>0.6) were used to annotate all shared loci, instruments, coloc regions, and SMR genes, and to rerun bidirectional MR after excluding inversion proxies (conclusion unchanged; OA → PD harmonized instrument sets contained no 17q21 instruments; PD → OA results were unchanged after removing rs199452). Stratification by haplotype was not possible at the summary-statistics level.

#### SMR + HEIDI

Summary-based MR (SMR)^43^ was used to test gene-expression–trait associations at the gene level, using brain eQTL (MetaBrain bulk^24^; basal ganglia and cortex) with blood eQTL (eQTLGen^44^) as a non-specific control. HEIDI was used to filter LD-driven false positives (p thresholds 0.05/0.01 reported). SMR results were cross-checked against SuSiE-coloc^17^; gene-level candidates (SMR significant + HEIDI passing + coloc-consistent) were prioritized into downstream cell-type-specific and expression analyses.

#### MVMR mediation (genetic mirror)

Two-step multivariate MR^25^ tested genetic mediation: step 1, OA → mediator (chronic pain, depression, insomnia, physical activity) with conditional F reported; step 2, OA + mediator → PD, computing indirect effects and proportion mediated with Delta-method CIs. BMI was included as a common covariate. Results were compared with the phenotypic mediation direction; discrepancies were reported (a weak genetic mediation does not falsify a behavioural/symptom-pathway explanation). Per the frozen decision rule, if the step-2 conditional F<10 (expected 24/24), indirect effects are described as “evidence insufficient (power-limited)” rather than “no effect,” and no mediation proportion is reported; the step-1 OA → pain instruments were adequate (conditional F 11.8–30.9).

### Cell-type-specific analyses

#### eQTL data and instruments

Cell-type-specific cis-eQTLs were obtained from Bryois 2022^12^ (eight brain cell types from frontal/temporal cortex and white matter; n = 192 donors; snRNA-seq; Zenodo 5543734/7276971), restricted to cis windows of gene TSS ±1 Mb, MAF>0.01, and INFO≥0.4. Two instrument sets were retained per (cell type × gene): (i) the lead eQTL SNP and (ii) all significant cis-eQTLs (FDR<0.05). Alleles were harmonized and strand-checked against 1000G EUR. We explicitly note that Bryois 2022 does not include midbrain dopaminergic neurons; neuronal evidence represents cortical excitatory/inhibitory neurons, and no claims about dopaminergic-neuron eQTLs are made.

OA-side instruments were the genome-wide significant independent loci (P<5×10⁻⁸; clumped at rZ<0.001, 10 Mb; F>10; MHC excluded). An eQTL SNP that also appeared in OA or PD loci was not removed a priori but adjudicated by colocalization in the cell-type-specific layer.

#### Cell-type-specific MR (csMR)

csMR followed a two-step design: (1) gene → PD MR for each (cell type × gene) using cis-eQTL instruments, with IVW (random effects) as primary and weighted median, MR-Egger, and weighted mode as sensitivity (Wald ratio for single-SNP genes); (2) OA × eQTL colocalization in each lead-eQTL region as a mediation premise. A candidate mediator required: (i) gene → PD MR FDR (BH) <0.05 within the cell type with a stable direction; (ii) OA × eQTL coloc PP.H4>0.8 (0.5–0.8 suggestive); and (iii) three-way direction consistency (OA risk allele × eQTL effect × MR direction). Formal criteria prioritized multi-instrument genes (N_IV≥2); single-SNP results were exploratory and additionally required coloc PP.H4>0.8, leave-one-out, and Steiger consistency. Shared-instrument matrices were reported when the same SNP appeared across cell types, to avoid spurious cell-type specificity.

#### ccFDR and annotation

Conditional/conjunctional FDR was computed for each (OA subtype × PD) pair on the harmonized SNP set, with conjFDR<0.05 primary and <0.01 strict, deduplicated at rZ<0.1, and annotated (FUMA/ANNOVAR: nearest gene, functional category, eQTL overlap).

#### Cell-type enrichment of shared loci

ConjFDR-significant loci were tested for overlap with each cell type’s eQTL loci using Fisher’s exact test plus 1,000 permutations with MAF/gene-density-matched random SNPs; cell-type specificity indices (tau; maximum-effect cell type) were used for secondary ranking; bulk eQTL (MetaBrain^24^), blood eQTL (eQTLGen^44^), and matched random loci served as controls. Results are reported as cell type × enrichment OR/95%CI/P (BH-corrected).

#### Single-cell disease-relevance scoring (scDRS), EWCE, and MAGMA

scDRS^45^ used OA (knee/hip/all) and PD GWAS summary statistics to compute per-cell disease scores in PD midbrain snRNA-seq data (dopaminergic neuron subtypes, microglia, astrocytes, etc.), testing OA-signal enrichment against PD signals, random gene sets, and permutations. EWCE^46^ and MAGMA-celltype^47^ provided orthogonal gene-level validation against cell-type marker genes. Multiple testing was controlled by cell type × method FDR; results were stratified into pre-specified priority cell types and whole-cell-type scans.

#### Three-trait colocalization

Three-trait coloc-SuSiE was performed in regions of csMR candidate genes and ccFDR shared loci (±500 kb), first as pairwise (OA × eQTL, eQTL × PD, OA × PD) and then integrated across the three traits. PP.H4>0.8 defined strong colocalization; 0.5–0.8 suggestive; PP.H3 and H3/H4 combinations were reported, with single-versus multi-signal sensitivity.

#### Sensitivity analyses

OA subtype consistency (knee/hip/all); PD replacement (Nalls 2019^20^ → Kim 2024^23^); eQTL threshold (FDR<0.05 vs P<1×10⁻⁵); MHC exclusion before/after; Steiger direction filtering; MR-Egger pleiotropy; and cross-checking against expression-validation direction consistency. When expression validation was negative and cell-type-specific analyses positive, the conclusion was downgraded to “shared but expression not validated.”

#### Functional enrichment analyses

Gene-set and tissue-expression analyses were performed with MAGMA v1.10^47^ on the gene-level results described above (35 kb/10 kb annotation window, 1000 Genomes Phase 3 EUR reference; six traits: FinnGen knee/hip/all OA, Hatzikotoulas all OA, Nalls 2019 PD, IPDGC no-UKBB PD). Gene-set analysis used the MSigDB v7.4^48^ C5 GO (biological process, cellular component, molecular function) and C2 canonical KEGG collections (10,371 sets in total), with one-sided competitive tests and BH-FDR per trait. Drug-target enrichment used DGIdb v5.0.11 (December 2023)^49^: a druggable-gene set (≥1 drug interaction), an approved-drug-target set, and 26 gene-category sets (≥20 mapped genes), analysed with the same MAGMA procedure. Tissue-expression analysis used the GTEx v8 54-tissue average log2-TPM matrix^50^ (MAGMA-ready file; Ensembl gene identifiers converted to Entrez via NCBI37.3 annotations) with the MAGMA gene-property model, testing each tissue separately with BH-FDR per trait. Candidate-gene enrichment used one-sided Fisher exact tests of the 141 csMR FDR-significant genes (union across PD outcomes) and the 68 evidence-supported subset against the GO/KEGG and DGIdb sets, with the 17,942 MAGMA-tested genes as background and BH-FDR across tests per candidate set. These analyses were exploratory and are recorded as a documented amendment (Supplementary Note N4); full results are provided in Fig. 8 and Supplementary Tables S13–S15, and the analysis scripts are archived in the OSF project.

### Single-cell expression validation

Expression validation used three independent PD midbrain snRNA-seq datasets: Kamath 2022^15^ (dopaminergic-neuron vulnerability map), Smajić 2022^14^ (glial activation and PD-specific neuronal states), and Martirosyan 2024^16^; datasets containing PD cases and controls from the substantia nigra were prioritized. Candidate gene lists were frozen from: cell-type-specific candidates (conj SNP-mapped genes plus csMR/coloc-positive genes); literature-anchored genes (ARL17A/ARL17B, KANSL1, WDR43, per Gu 2026^13^); and OA-side risk genes mapped from genome-wide significant OA loci.

Differential expression was estimated with pseudobulk DESeq2 as the primary approach (cells aggregated within cell type and donor; the primary model included age, with sex and post-mortem interval added in separate sensitivity models; batch covariates were not available), with single-cell-level MAST as a secondary approach. Vulnerable dopaminergic subtypes (e.g., SOX6+/ALDH1A1−) were stratified and reported separately. Multiple testing was controlled by cell type × gene BH-FDR. Output included cell type × gene expression heatmaps and effect-direction consistency tables against csMR/single-cell enrichment.

### Statistical thresholds and multiple testing

Multiple-testing corrections were applied per analysis layer with pre-specified thresholds, as summarised in Table 6. Effects are reported as OR/β with 95% CIs and P values; strata with fewer than 10 events are reported for direction only, with power limitations stated.

**Table 6.** Multiple-testing strategy by analysis layer.

| Layer | Multiple-testing strategy |
| --- | --- |
| Phenotypic | 3 mediator paths (NHANES); specificity across 4 exposure categories → BH-FDR; E-values |

|  | reported |
| --- | --- |
| LDSC rg | Bonferroni across combinations × OA subtypes × PD versions; 95% CI reported |
| MR | 4 estimators per direction; primary judgment = IVW and weighted-median consistency; Egger intercept as pleiotropy test |
| ccFDR | conjFDR<0.05 primary, <0.01 strict |
| LAVA | FDR over local windows |
| scDRS/EWCE/MAGMA-celltype | FDR across cell types × methods |
| csMR | FDR across cell types; coloc PP.H4>0.8 |
| Expression | BH-FDR across cell types × genes |
| Functional enrichment (MAGMA gene-set, DGIdb, GTEx) | One-sided competitive/gene-property tests; BH-FDR per trait within each test family (gene sets; DGIdb categories; 54 tissues); six traits tested |
| Candidate-gene enrichment (141 csMR genes; 68 evidence-supported) | One-sided Fisher exact tests vs GO/KEGG/DGIdb sets; BH-FDR across tests per candidate set (background: 17,942 genes) |

### Software and reproducibility

Analyses used Python 3.12 (numpy/scipy/pandas/statsmodels); R 4.6.1 with coloc 5.2.3, susieR, data.table, survey, mediation, sandwich, clubSandwich, TwoSampleMR, and MVMR; PLINK 1.9 (clumping rZ=0.001/10 Mb); LDSC 2.0.1 (with a documented allele-filtering patch); MAGMA v1.10^47^; and the standard conditional-FDR estimator (Python) for ccFDR. All intermediate files (munged summary statistics, clumping outputs, logs) are retained with SHA records; the full pipeline and version manifest accompany the submission. Scripts are archived in the OSF project. Full software and package versions are listed in Supplementary Note N5.

## Data Availability

All data produced in the present study are available upon reasonable request to the authors.

## Data availability

All data sources, accessions, versions, and md5 checksums are listed in sources_manifest.csv and Supplementary Table S1. The OSF project (https://osf.io/rgeaf) contains the analysis protocols, data dictionaries, and archived results, together with the OSF registration (completed 25 August 2026; DOI 10.17605/OSF.IO/K8Q5A). All datasets were used under their respective public data-use agreements (FinnGen, UK Biobank–derived summary statistics, CHARLS, NHANES, IPDGC/GP2, eQTLGen, MetaBrain, GTEx, Zenodo); access conditions are recorded in sources_manifest.csv and Supplementary Table S1. Analyses were run on the v2 pipeline before the OSF registration and re-run on the archived v3 pipeline after the retrospective registration as a verification comparison; v2–v3 differences were limited to weighted-median/weighted-mode bootstrap SE and P, with point estimates identical across all compared values and conclusions unchanged (comparison archived in the OSF project).

## Supplementary information

Table S1 data sources; S2 baseline characteristics; S3 phenotypic models/sensitivity; S4 LDSC/LAVA; S5 bidirectional MR full (including combination D instrument-set replication); S6 overlap matrix/gcov; S7 ccFDR full; S8 coloc full; S9 SMR/HEIDI; S10 MVMR; S11 csMR/scDRS/EWCE; S12 expression; S13 GO/KEGG pathway enrichment; S14 DGIdb drug-target enrichment; S15 GTEx tissue enrichment; Supplementary Fig. 1 technical route of the study; S2 CHARLS sensitivity; S3 NHANES sensitivity; S4 EWCE cell-type enrichment; S5 csMR sensitivity forest plot; S6 shared OA–PD instruments; S7 ccFDR Manhattan; S8 three-way direction heatmap; S9 MAGMA gene-level volcano plot; S10 pseudobulk DESeq2 differential expression. Supplementary Notes: N1 OA × AD contrast; N2 MAPT 17q21 H1/H2 inversion sensitivity; N3 CAUSE analysis declaration; N4 documented amendments; N5 software versions; N6 forest-plot annotations. Supplementary Checklists: STROBE; STROBE-MR.

## Code availability

Analysis scripts are archived in OSF component 09_Code, and the executed result tables, logs, and reproducibility records are in component 10_ModelsRun. All analyses were run on the archived v3 pipeline under a pinned TwoSampleMR version with random-effects IVW and set.seed(2026), as documented in the OSF project.

## Supplementary figures

**Supplementary Fig. 1.**
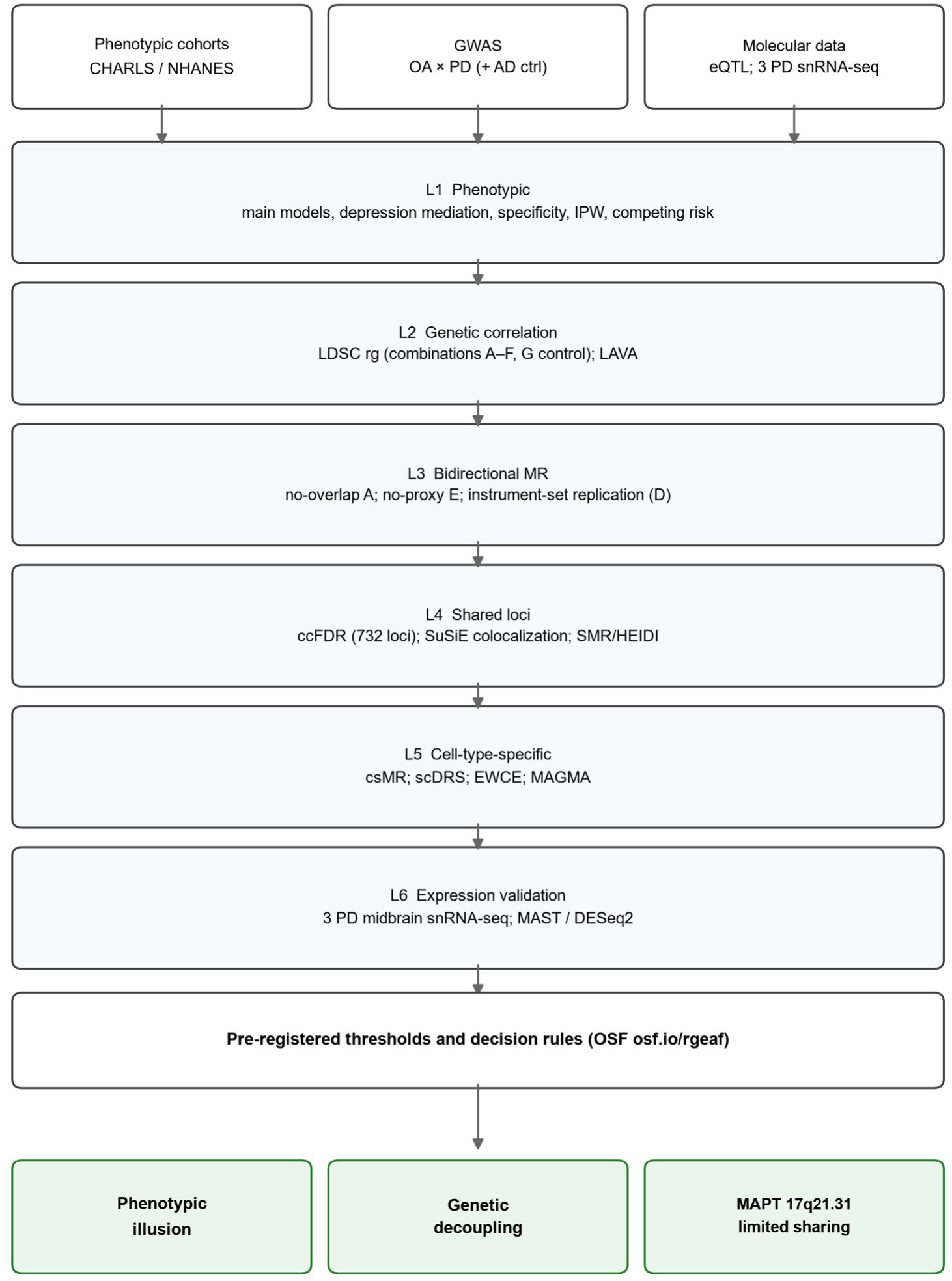
Technical route of the study. Phenotypic cohorts (CHARLS, NHANES), OA/PD GWAS (FinnGen, Hatzikotoulas, Nalls, IPDGC no-UKBB, Kim 2024, with a Bellenguez AD control), brain cell-type eQTLs (Bryois 2022; MetaBrain/eQTLGen), and three PD midbrain snRNA-seq datasets feed six analysis layers (phenotypic mediation and specificity; LDSC/LAVA genetic correlation; bidirectional MR with combination-D replication; ccFDR/SuSiE-coloc/SMR-HEIDI shared-locus mapping; csMR, scDRS, EWCE and MAGMA cell-type analyses; expression validation). Thresholds and decision rules pre-specified in the analysis protocol govern the final interpretation (phenotypic illusion; genome-wide genetic decoupling; limited direction-unbiased sharing at MAPT 17q21.31).

**Supplementary Fig. 2.**
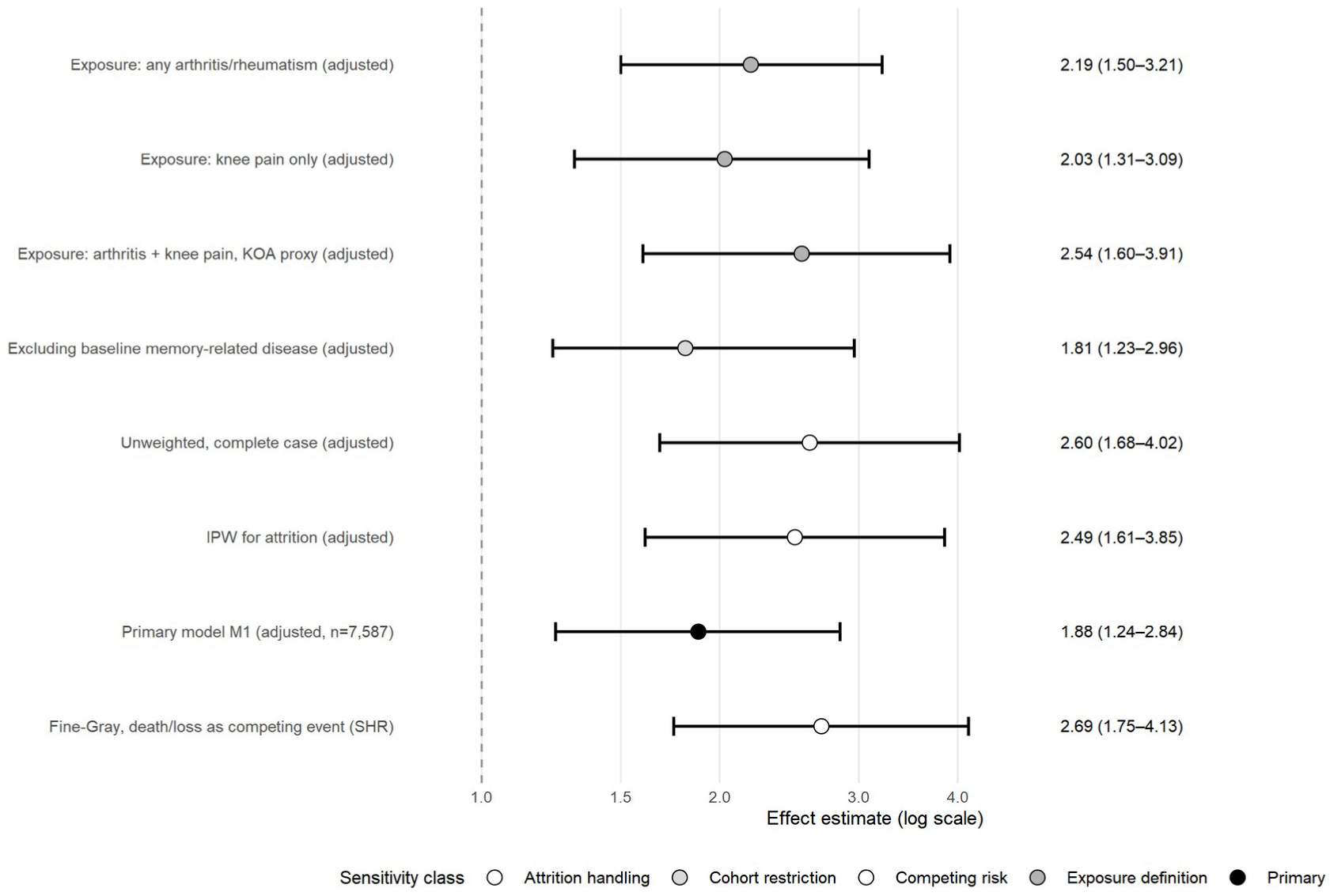
CHARLS sensitivity analyses (v3). All models are adjusted; each row uses its own pre-specified sample/definition (primary: n = 7,587, excluding baseline stroke/memory disease; exposure-definition rows: larger sample without baseline exclusion; unweighted/IPW: complete-case 2020-follow-up sample, same model with and without attrition inverse-probability weights; Fine-Gray: subdistribution hazard ratio with death/loss as competing event). Point estimates are directionally consistent and significant across all specifications; within-pair comparisons (unweighted vs IPW) are directly comparable, whereas cross-group differences reflect definition/sample differences.

**Supplementary Fig. 3.**
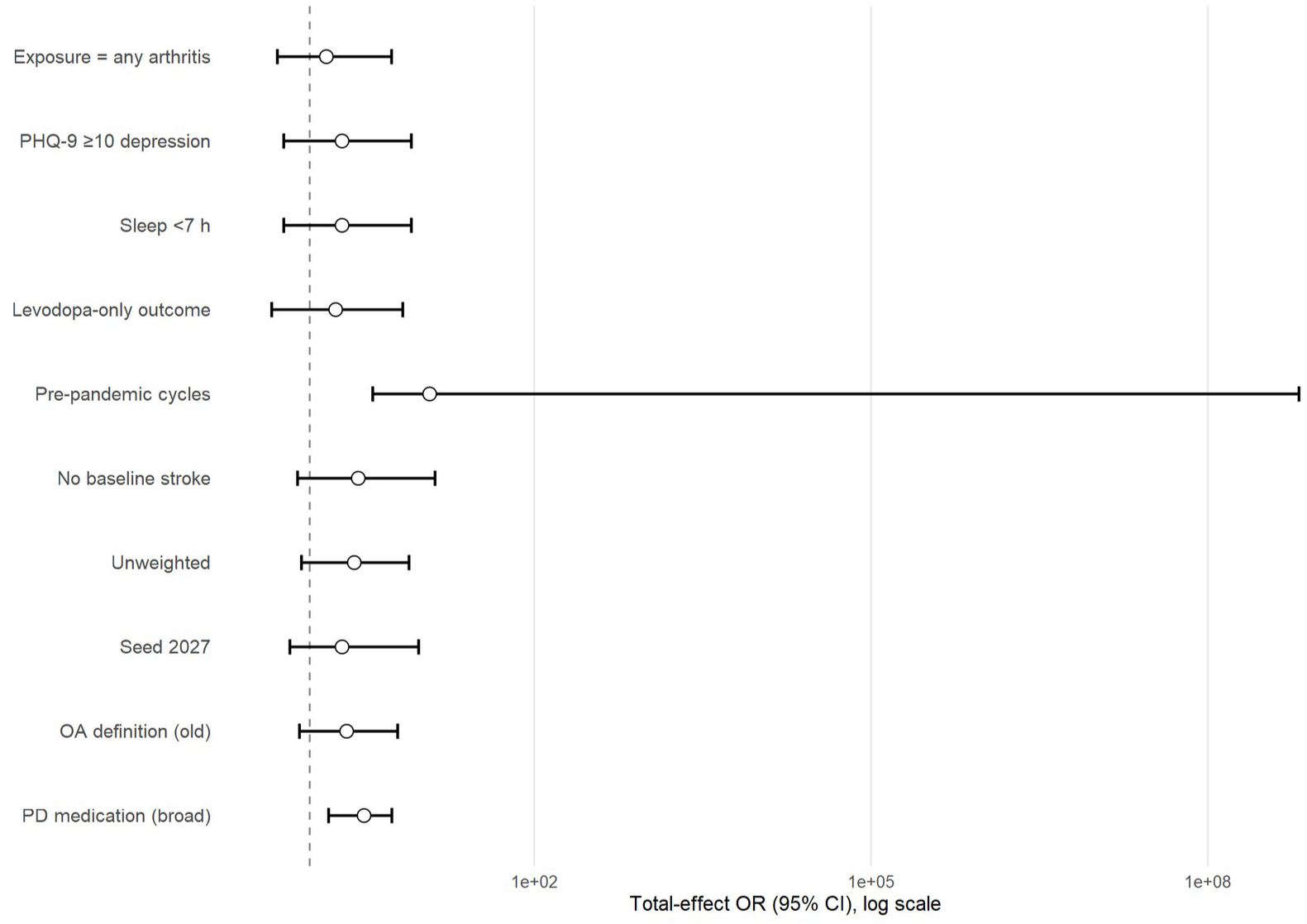
NHANES sensitivity analysis (v3). Total-effect ORs (95% CI) for the OA–PD association across the ten pre-specified sensitivity scenarios (exposure definition, outcome definition, weighting, pre-pandemic P-file standalone, and random-seed checks).

**Supplementary Fig. 4.**
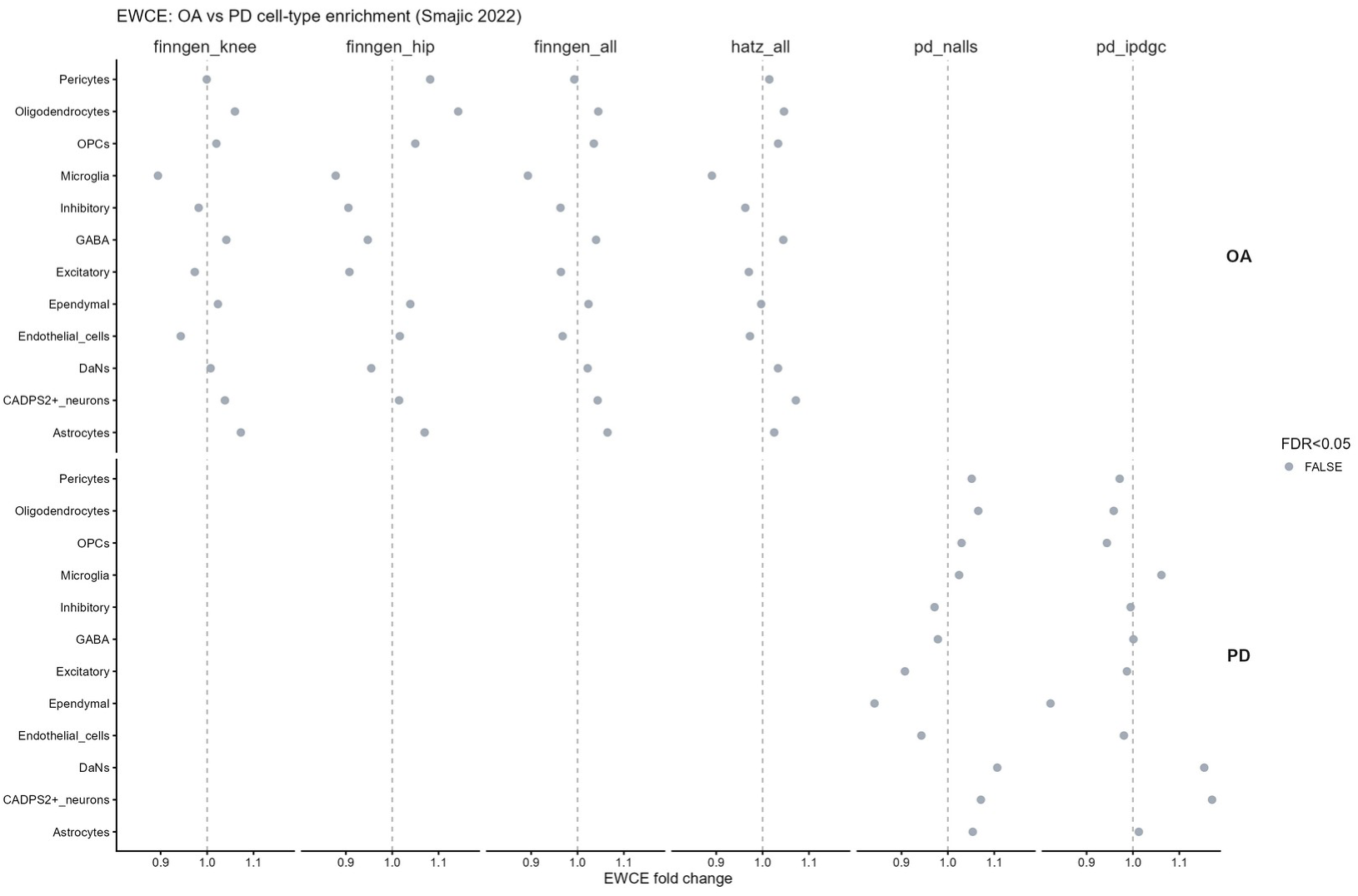
EWCE cell-type enrichment for OA and PD shared loci.

**Supplementary Fig. 5.**
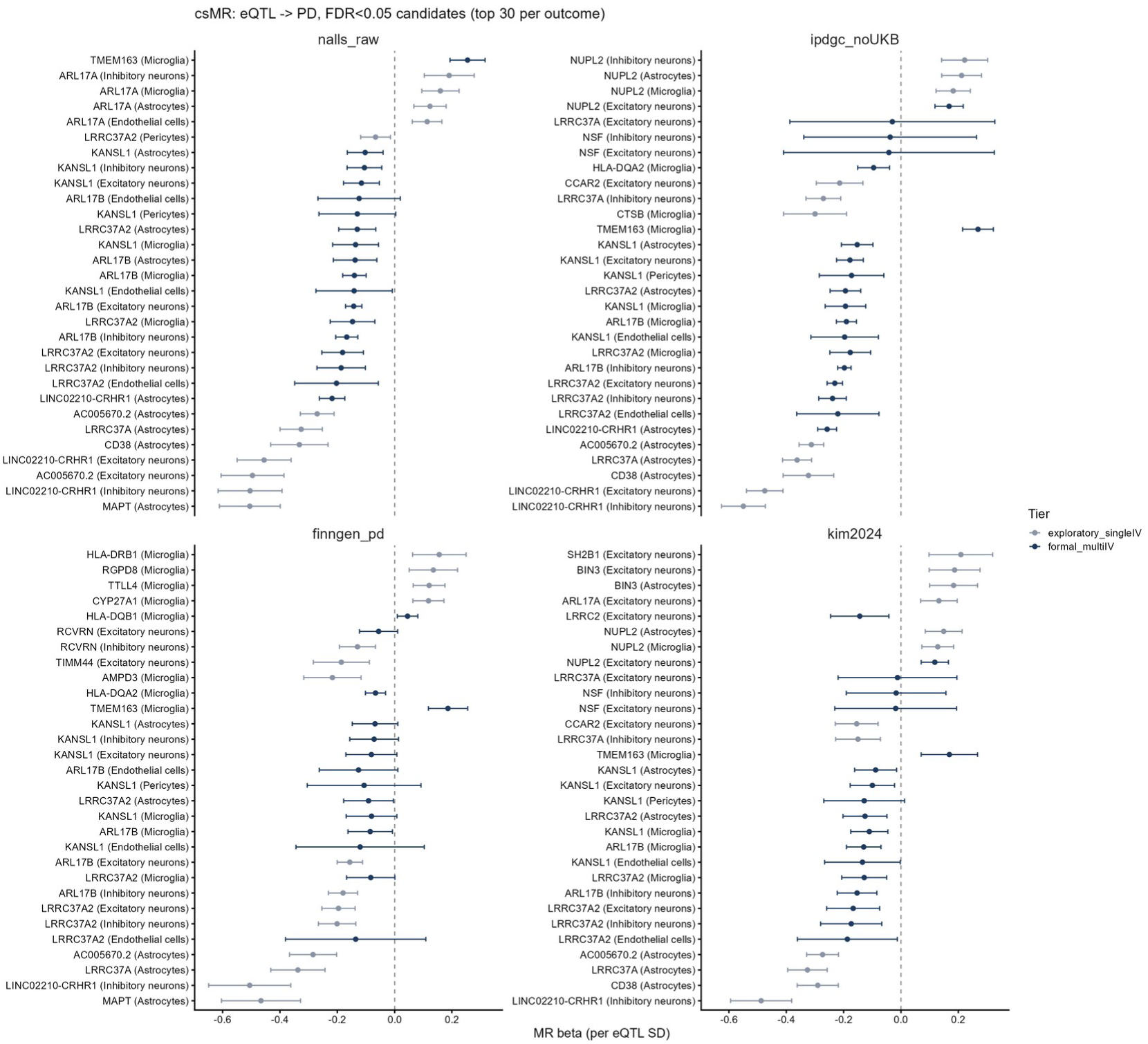
csMR sensitivity forest plot. Gene–PD cell-type-specific MR estimates with sensitivity estimators across the four PD GWAS.

**Supplementary Fig. 6.**
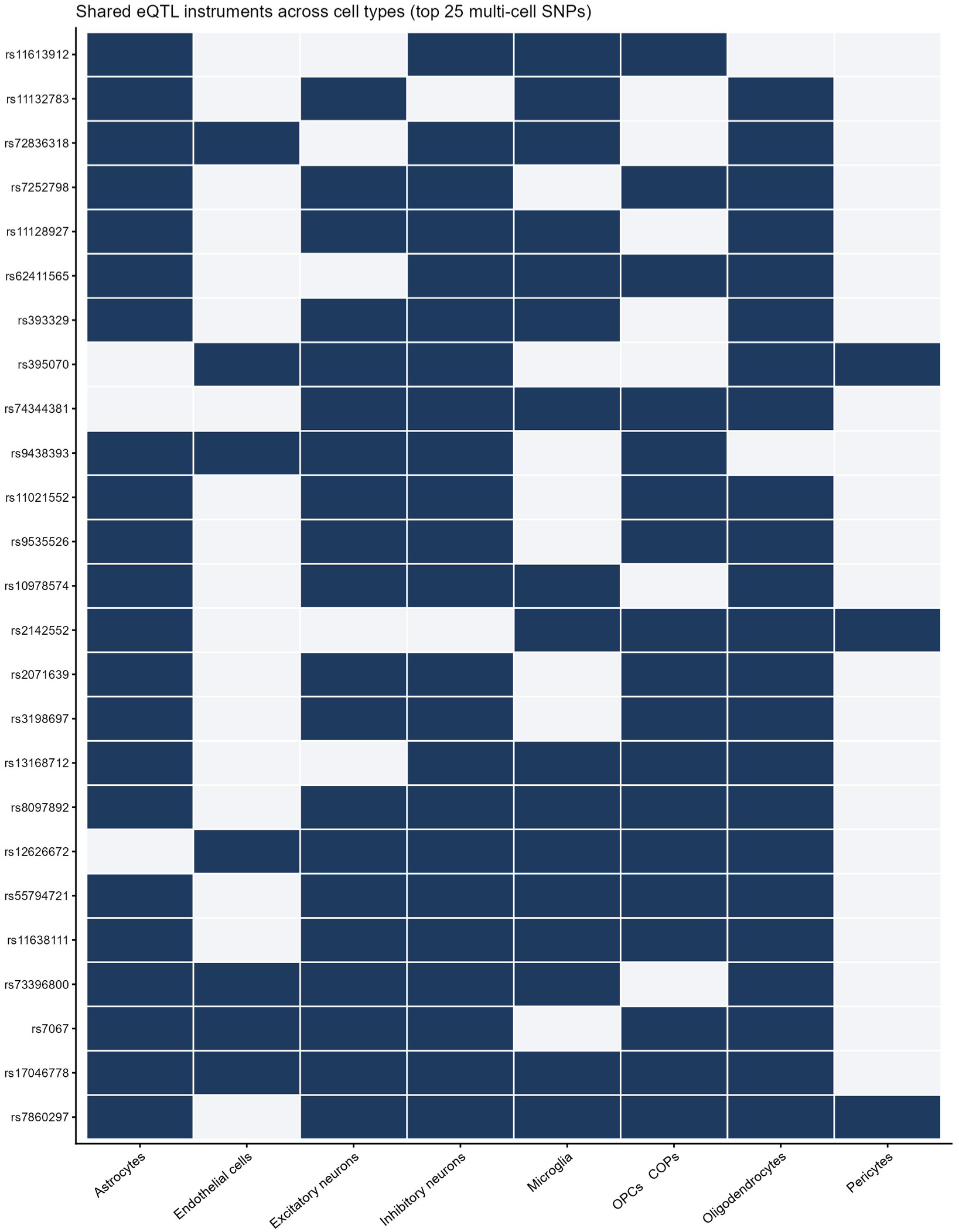
Shared OA–PD instruments across combinations. Overlap of instruments used in bidirectional MR and ccFDR shared-locus mapping.

**Supplementary Fig. 7.**
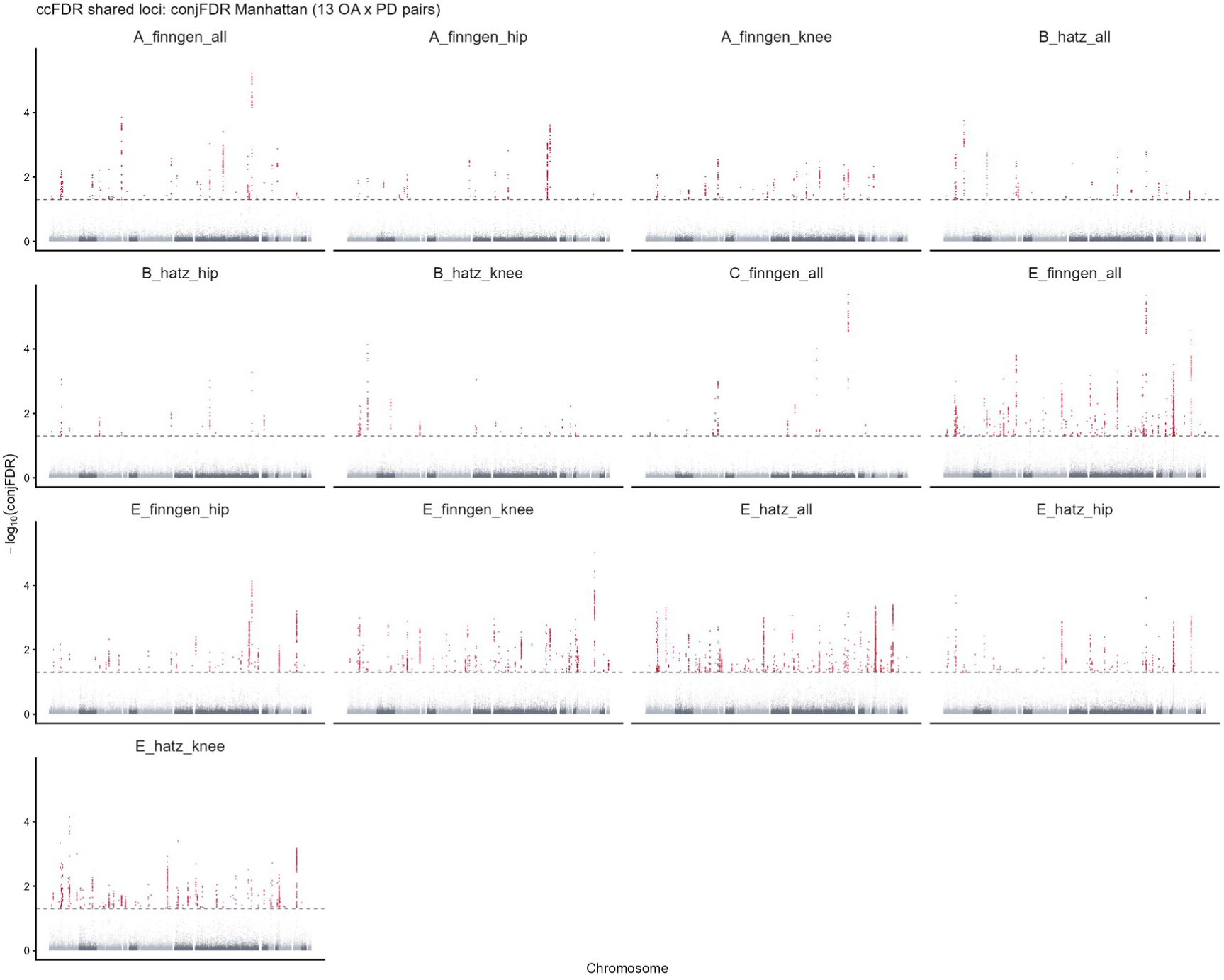
ccFDR Manhattan across the 13 OA × PD pairs. conjFDR<0.05 highlighted; clumped shared loci used for directional analysis (main Fig. 5a).

**Supplementary Fig. 8.**
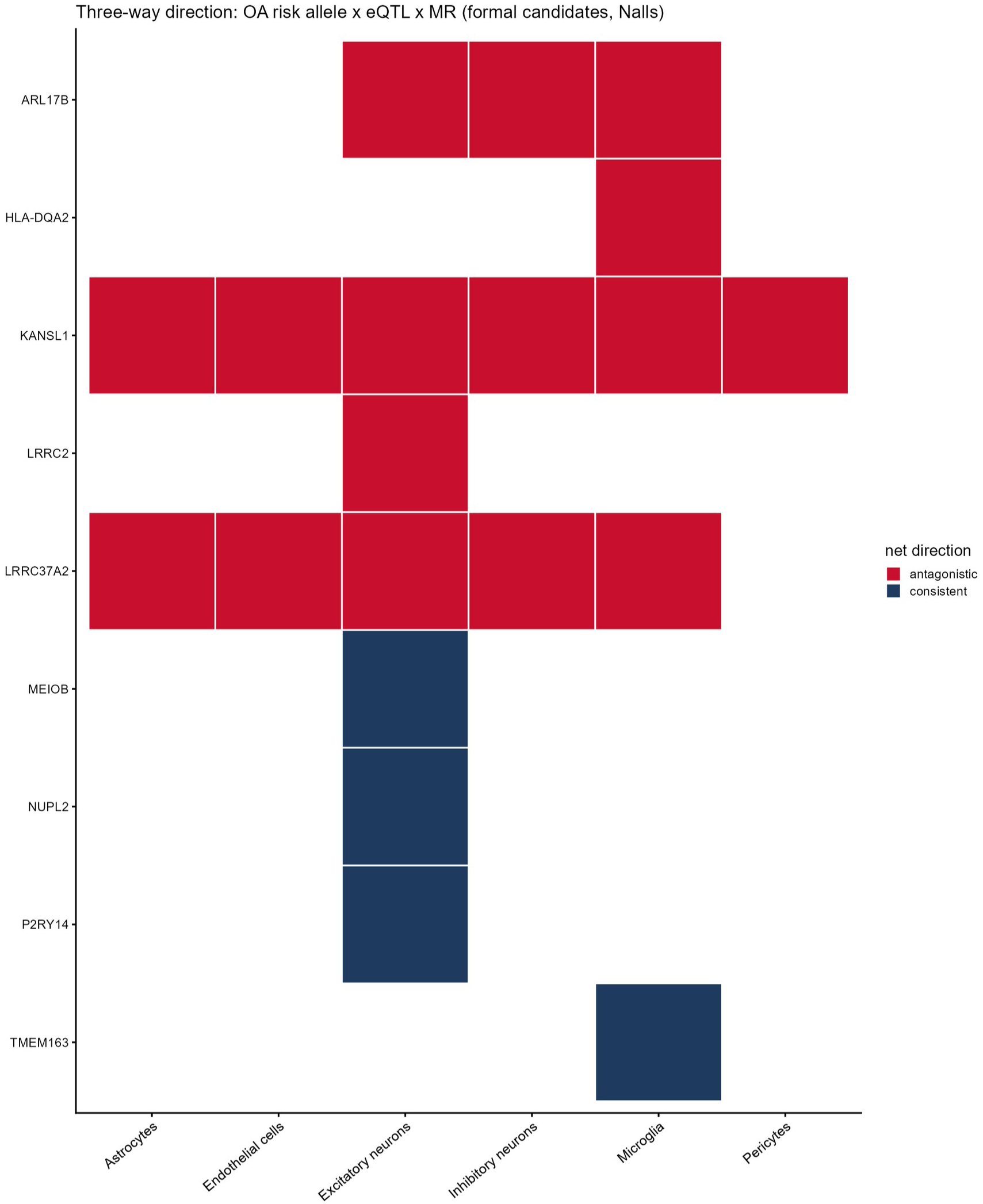
Three-way direction heatmap. OA risk allele × eQTL × MR net chain for formal csMR candidates (blue = consistent, red = antagonistic; Nalls outcome).

**Supplementary Fig. 9.**
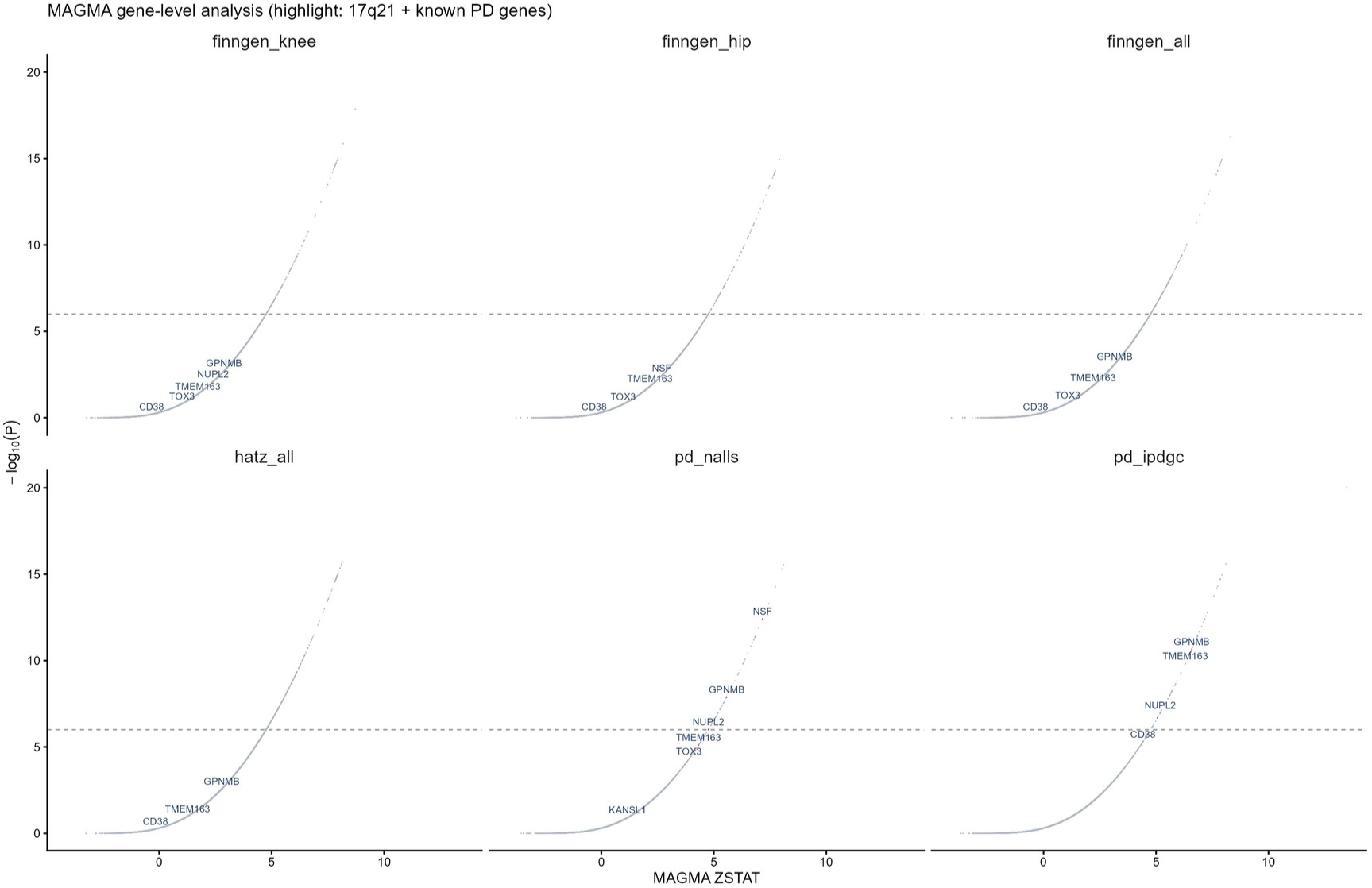
MAGMA gene-level volcano plot. 17q21 and known PD genes highlighted.

**Supplementary Fig. 10.**
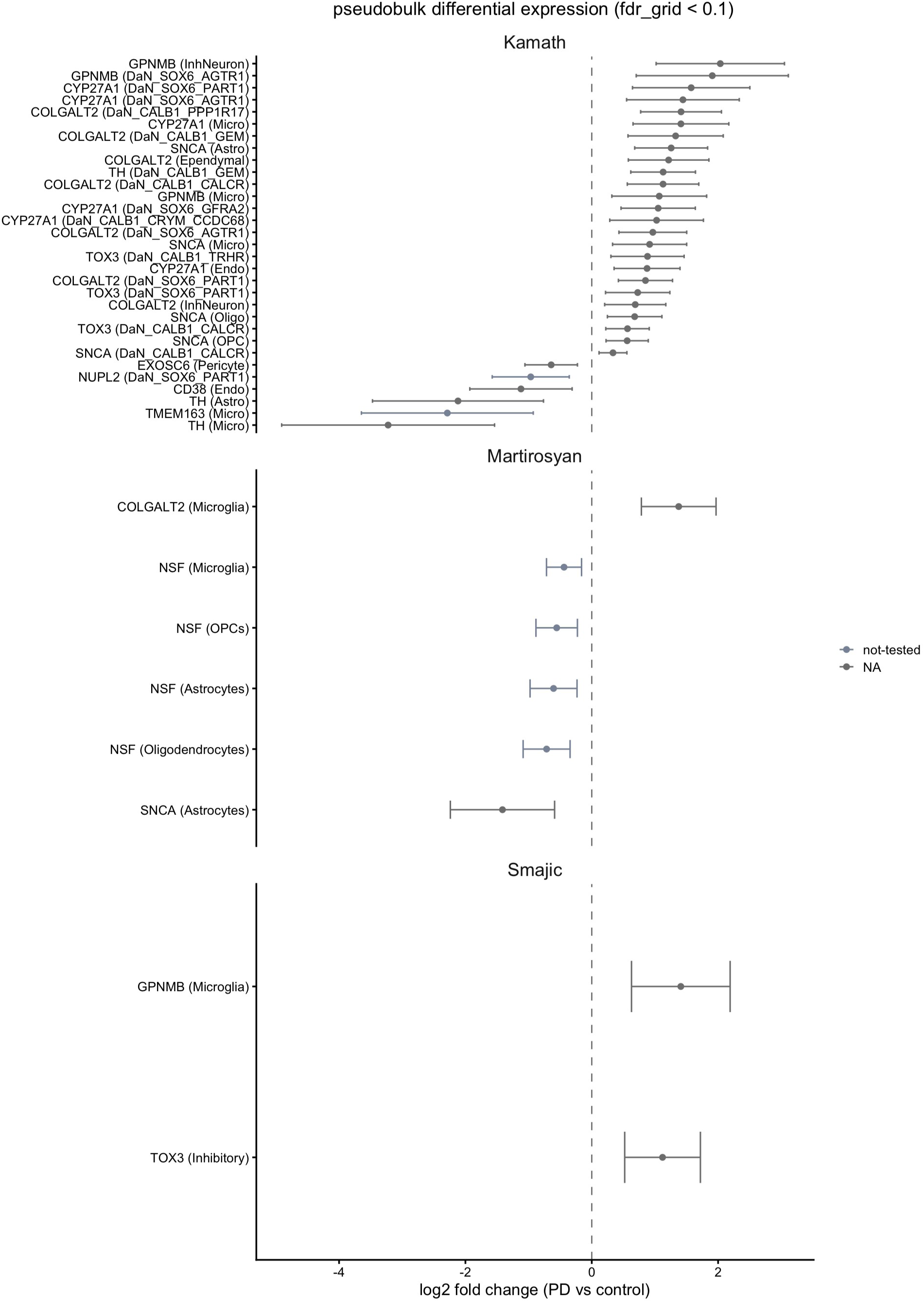
Pseudobulk DESeq2 differential expression. Significant gene × cell-type pairs (padj<0.05) with log2 fold change and 95% CIs; Smajić, Martirosyan, and Kamath datasets.

## Declarations

### Author contributions

(CRediT): S.Y. conceived and designed the study and wrote the original draft; S.Y., Z.W. and H.Z. performed the statistical and genetic analyses, curated the data and developed the software; S.Y., Z.W., H.Z. and P.T. contributed to interpretation and validation of results; Z.W., H.Z. and P.T. reviewed and edited the manuscript; P.T. supervised the study. All authors read and approved the final manuscript. Competing interests: the authors declare no competing interests.

## Acknowledgements

We thank the China Health and Retirement Longitudinal Study (CHARLS), the National Health and Nutrition Examination Survey (NHANES), FinnGen, the International Parkinson Disease Genomics Consortium (IPDGC), the Global Parkinson’s Genetics Program (GP2), the GWAS Catalog, eQTLGen, MetaBrain, the Genotype-Tissue Expression (GTEx) Project and Zenodo for making their data publicly available.

